# Spatially resolved simulations of the spread of COVID-19 in European countries

**DOI:** 10.1101/2020.11.25.20238600

**Authors:** Andrea Parisi, Samuel P. C. Brand, Joe Hilton, Rabia Aziza, Matt Keeling, D. James Nokes

**Affiliations:** School of Life Sciences and Zeeman Institute for Systems Biology and Infectious Disease Epidemiology Research (SBIDER), University of Warwick, UK; Kenya Medical Research Institute (KEMRI) - Wellcome Trust Research Programme, Kilifi, Kenya

## Abstract

We explore the spatial and temporal spread of the novel SARS-CoV-2 virus under containment measures in three European countries based on fits to data of the early outbreak. Using data from Spain and Italy, we estimate an age dependent infection fatality ratio for SARS-CoV-2, as well as risks of hospitalization and intensive care admission. We use them in a model that simulates the dynamics of the virus using an age structured, spatially detailed agent based approach, that explicitly incorporates governamental interventions, changes in mobility and contact patterns occurred during the COVID-19 outbreak in each country. Our simulations reproduce several of the features of its spatio-temporal spread in the three countries studied. They show that containment measures combined with high density are responsible for the containment of cases within densely populated areas, and that spread to less densely populated areas occurred during the late stages of the first wave. The capability to reproduce observed features of the spatio-temporal dynamics of SARS-CoV-2 makes this model a potential candidate for forecasting the dynamics of SARS-CoV-2 in other settings, and we recommend its application in low and lower-middle countries which remain understudied.

## 1 Introduction

The first cases of COVID-19, caused by the novel severe acute respiratory syndrome coronavirus (SARS-CoV-2), were reported in Wuhan, China in December 2019. Rapidly causing a local epidemic, it spread around the world within a few months causing a major pandemic which by the start of May 2020 had resulted in over 1,000,000 cases and over 300,000 deaths worldwide. Early data from China suggested levels of transmissibility and virulence comparable to those observed in the Spanish flu epidemic of 1918-19, which caused an estimated 20-50 million deaths worldwide [1]. Despite causing mild infections in the majority of cases, a small but considerable fraction of infected individuals requires hospitalization and, in the most severe cases, intensive care with mechanical ventilation. Estimates of the case fatality ratio (CFR) show a dramatic variation across populations [2], likely reflecting the diverse approaches to case detection adopted by different countries. In the absence of an effective vaccine able to fend off the infection, many countries have adopted stringent non-pharmaceutical control measures designed to reduce transmission rates by limiting the frequency of person-to-person contact. These measures are designed to limit the total mortality due to COVID-19 and to confine the epidemic to a level that does not overwhelm national health systems.

As a novel infectious agent, many characteristics of SARS-CoV-2 are still poorly understood and are the object of close scrutiny by the research community worldwide. Early estimates of the basic reproductive number in Hubei province, China, varied between 2.2 [3] and 3.11 [4], with estimates in European countries ranging between 2.5 and 3.0 [5, 6, 7]. Measurements of the doubling time ranged between 2.9 and 7.4 days [5, 3]. The majority of infections are asymptomatic, but there is uncertainty over the proportion of asymptomatic infections, with reports ranging between 4.5% and 64% [8, 9, 10, 11]. A study conducted in the municipality of Vo’ Euganeo, at the centre of the Italian outbreak, in which the whole population was tested irrespective of symptoms across two consecutive surveys found the mean proportion of asymptomatics to be between 40-45%[10]. An outbreak onboard the cruise ship Diamond Princess led to most passengers and staff tested for COVID-19, with 51% of laboratory confirmed cases being asymptomatic [12] and a subsequent modelling study estimating the asymptomatic proportion 74% (95% C.I. 70-78%) [13]. The proportion of cases which are asymptomatic is of interest because of their role in the transmission of the virus within the population. However, so far age stratified fractions of asymptomatic cases remain difficult to obtain. Evidence from several studies [10, 14] suggest that children are less likely to be affected by coronavirus, and data from most countries show that the rate of severe cases in young children is low compared to the mature population, although it is still unclear whether this reflects lower susceptibility or higher rates of asymptomatic infection among children. A detailed survey conducted by the Spanish Government [15] showed that only 1% of children below 1 year of age were infected by the SARS-CoV-2 virus in Spain; this percentage increased with age to 2.2% for 1-4 years old children, and to higher percentages for older individuals, up to almost 7% for individuals aged 70-74. While this could reflect a lower susceptibility to the virus among young individuals, decoupling this variation from age-structured mixing effects requires a detailed mathematical analysis. Additionally, the Spanish survey showed that the number of infected individuals is substantially larger than the number of detected cases, suggesting that large numbers of undetected asymptomatic or mildly symptomatic cases may have driven the outbreak.

An age dependent fraction of infected individuals suffers from severe symptoms that require hospitalization, and a fraction of these require mechanical ventilation in intensive care units. Estimates of hospitalization rates based on Chinese data as well as the infection fatality ratios were estimated in Verity *et al*. [16]. Based on these, a set of estimates for hospitalization and admissions in intensive care were used by Ferguson *et al*. [17]. Recent data coming from the first highly infected European countries (Italy and Spain) provide space for further estimates. By combining recent information on cases, hospitalizations, intensive care admissions, deaths with results from the Spanish survey, we are able to build updated age stratified estimates of the ascertainment rate, rate of hospital admissions, intensive care admissions and corrisponding death rates.

A number of intervention were progressively implemented by most countries worldwide to counter the wide spread of the virus. These included stay-at-home measures, shielding of most-at-risk population, closure of schools and higher level educational institutions, teleworking, cancellation of mass gatherings, closure of shops, including recreational and food establishments [18]. The underlying idea of these interventions is to break the chain of transmission of the virus by reducing contact between individuals. In most countries, the interventions adopted resulted in a substantial reduction of the disease morbidity and mortality. The adoption of these intervention has typically been driven by intense modelling work carried out by national teams as well as the wider international research community. Analysis of big data provided by some large companies [19, 20] has been an important component of the modelling effort.

In this work, we present a detailed model of the spatial spread of SARS-CoV-2 virus under non-pharmaceutical interventions that explicitly incorporates governamental interventions, changes in mobility and contact patterns as implied by measurements by big data operators [19] as well as demographic, internal human mobility and age mixing information. Our aim is to have a model that could be readily used on a range of different countries, including middle and low income countries: we thus apply the model to three European countries, Italy, Spain and the United Kingdom, to assess its capability of properly reproducing observed data, and to obtain estimates of key parameters that are, as much as possible, country independent. We use data from the first serological survey performed in Spain [15] as well as data on Intensive Care Units (ICU) admissions and deaths in Italy [33] to infer rates of detection of cases, severe and critical admissions to hospital, as well as death rates due to SARS-CoV-2. We then use the model to estimate, for each country, an *unrestricted R*_0_, which is the value that the basic reproductive number would achieve in the absence of interventions. We project *R*_0_ to a reference country, China, thus removing the influence of country specific contact matrices. We also estimate a set of age-dependent susceptibilities independently for each country, resulting in a overall set of age stratified susceptibilities which agrees in all the countries under study. Output from our simulations is capable of reproducing the spatio-temporal spread of the disease in the studied countries, with some disagreement on the spatial aspect for Spain due to specific features of human mobility in that country.

## 2 Methods

The simulation program uses a gridded spatial description with a resolution of 5km. In each grid elements, age stratified estimates of the resident populations were obtained from the WorldPop database [21]. Maps available with a 100m resolution were coarsened to 5km: this choice was mainly due to computational efficiency. Each grid cell is treated as a well mixed population, whereas transmission between distinct grid elements is driven by human mobility. The model uses an agent-based description in which individuals have a set of preferred locations and an associated frequency of visit which are assigned at the start of the simulation. Each day individuals move to one of their preferred locations and participate to the local transmission dynamics. The list of preferred locations and their frequency of visit and duration of stay are built in accordance to the fluxes between grid elements predicted by an appropriate model for human mobility.

### 2.1 Epidemiological model

In each location, the dynamics follows a modified age stratified SEIR model that includes further classes for hospitalization (severe cases) and cases in intensive care (critical cases), for a total of 9 age groups and 11 distinct epidemiological classes: a schematic description of the model is shown in Fig. 1, while the full model is detailed in the Supplementary Material SM-1. The model distinguishes between two kind of infectives: detected cases *I*_*j*_ and undetected *U*_*j*_. Detected cases may evolve into severe or critical cases, but otherwise both categories have same infectiousness. All rates we derive may be expressed as rates per infected individual, regardless of its symptomaticity. We do distinguish between detected and undetected cases to provide corresponding estimates of prevalence.

**Figure 1:**
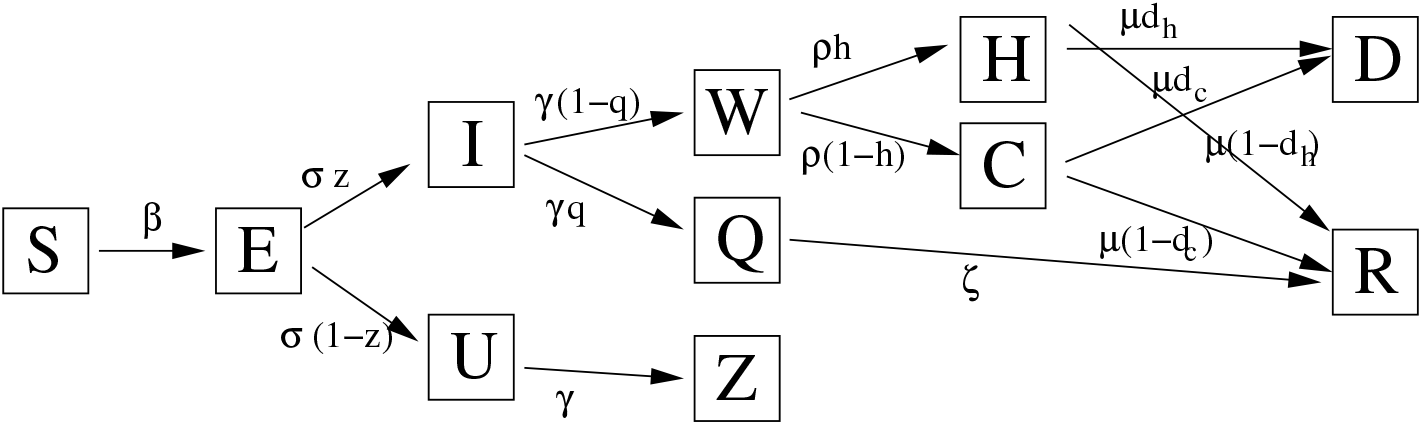
Schematic and simplified description of the epidemiological model. The full model is age-structured, and includes special handling of transmission in households as well as separating the transmission of work environments from other kind of transmission. S: susceptible; E: exposed; I: infective case; U: undetected case; H: hospitalized; C: critical; W: delay for hospitalised or critical; Q: quarantined at home; D: death; R: recovered; Z: removed asymptomatic.

**Figure 2:**
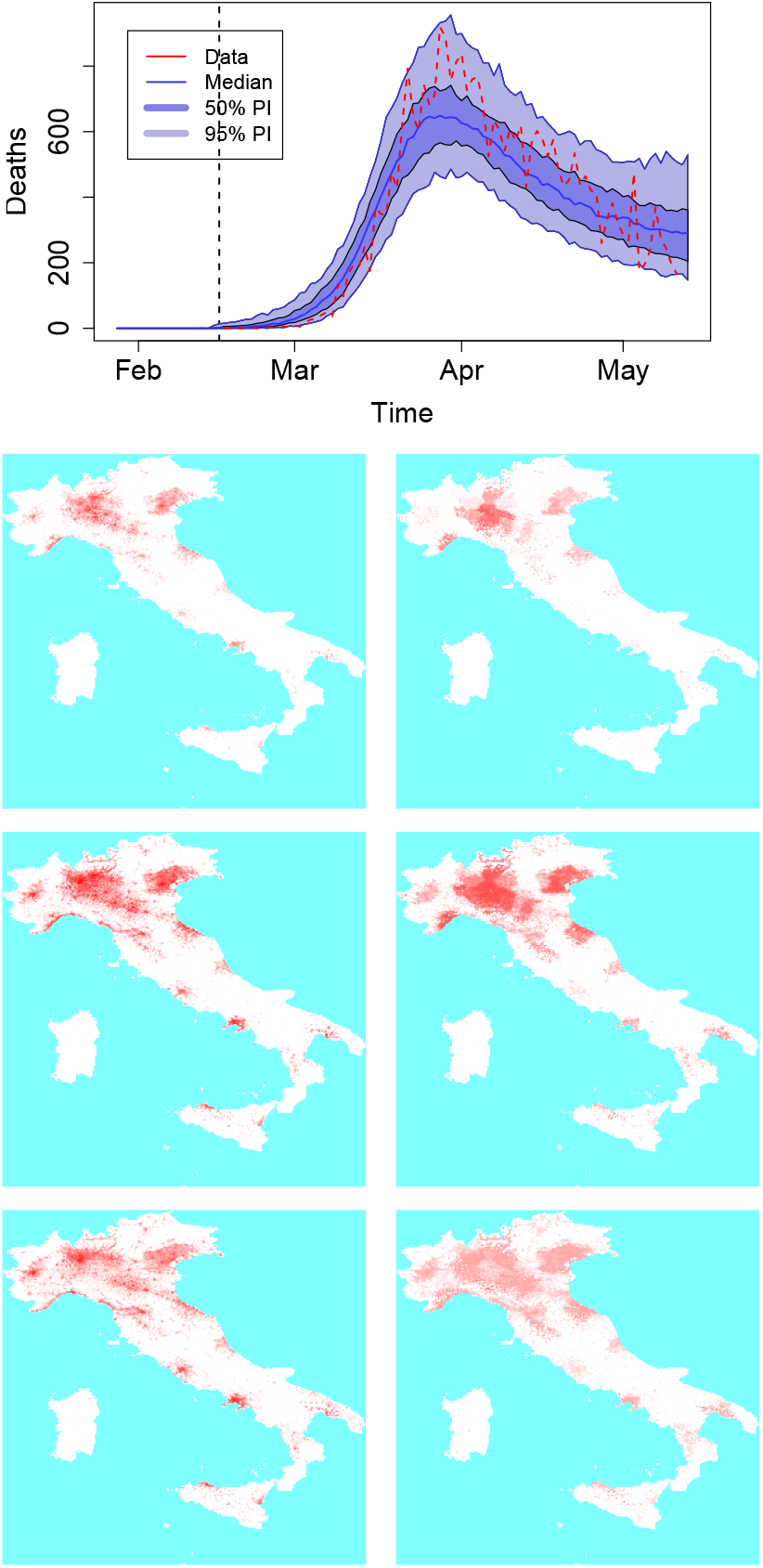
(Top) Country level dynamics of SARS-COV-2 inferred by death incidence data for Italy. Fitted parameters are *R*_0_ = 4.1 (95% CI [2.6 - 5.6]), *t*_0_ = 6 (95% CI [0 - 12]) days, *τ*_*γ*_ = 5.7 (95% CI [1.7-7.9]), and log *ω* = −7.5 (95% CI [−10 – −4.6]) days^−1^. (Bottom) Frames from an average of 10 simulation for Italy, corresponding to cases at the end of week 2, 4 and 7: is the average incidence, on the right the incidence as a fraction of the population.

**Figure 3:**
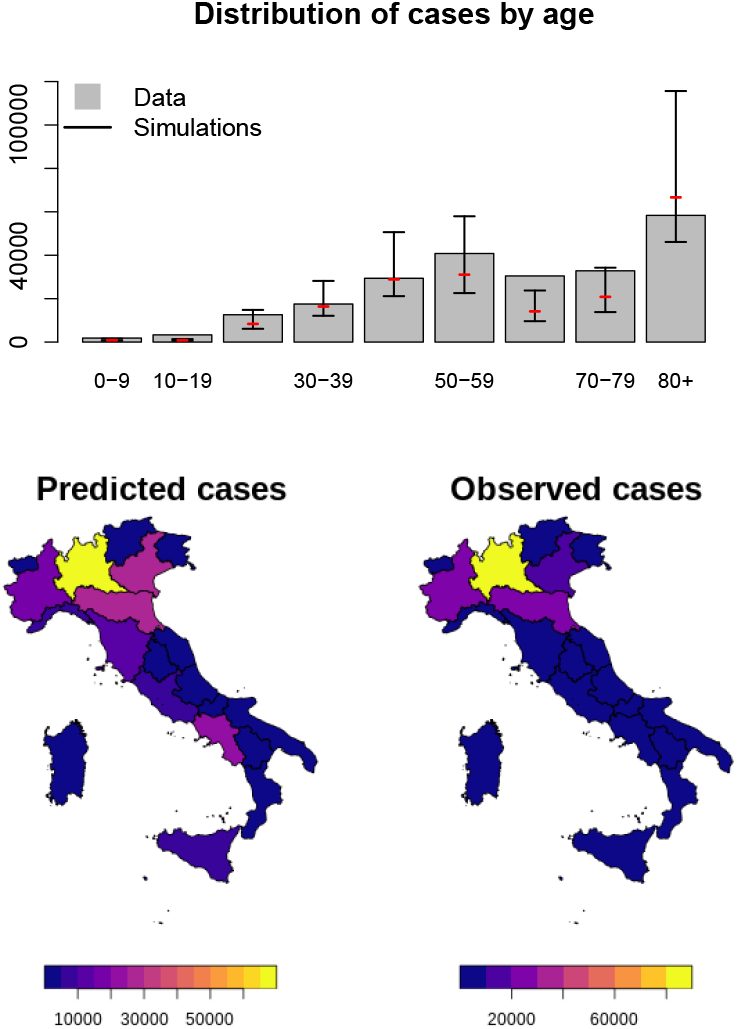
(Top) Age distribution of cases predicted by the model and comparison with data from Italy. The model overestimates cases in the middle age groups, and underestimates cases in the elderly. (Bottom) Comparison between predicted spatial spread of COVID-19 cases and observed distribution.

Transitions between compartments are driven by stochastic events: a direct Gillespie algorithm is used to integrate the stochastic dynamics, with the exception of transmission within households which are handled separately. Infections are driven by contacts between susceptibles and infectives, either detected or undetected. We use age-mixing patterns estimated by Prem *et al*. [22]: these consist of distinct estimated age-mixing matrices for home contacts, work, school and other kind of contacts. We take advantage of this subdivision to implement different control strategies. Given a susceptible in class *S*_*i*_, the force of infection is given by:

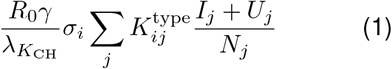

where *R*_0_ is the basic reproductive number for pre-lockdown China and 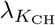 is the maximum eigenvalue of the Chinese contact matrix. This approach allows exporting estimates of the basic reproductive number obtained from Chinese data to other countries and was used previously in Brand *et al*. [23]. The age dependent susceptibilities *σ*_*i*_ are estimated from simulations. We consider work and non-work transmission separately, thus the type superscript may indicate one of the following two contact matrices: 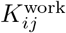 and 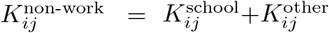 Due to this separation, we can implement stay-at-home interventions that affect one kind of transmission but not the other. The contribution from any of these matrices may be reduced or increased during the simulation.

### 2.2 Households

Home transmission is not included in the above, and is handled differently. Using contact matrices for home contacts in the context of an age stratified well mixed model, while reasonable during unconstrained transmission, risks to overestimate widespread transmission in presence of home confinement due to the absence of household structure. To overcome this issue, we include a description of households based on the dynamic generation of synthetic households. When an individual is infected, the simulation program will dynamically generate a household by linking together unrelated individuals. Suppose an individual in age group *j* not previously included in any household is infected, a host household is generated on-the-fly. First, a household size is chosen from a Poisson distribution with mean equal to the country mean household size. Then, susceptible individuals from age groups *i* are added to this household, randomly chosen according to the discrete probability distribution 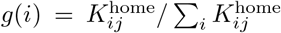. The choice of choosing susceptibles is driven by the fact that individuals from other classes would have already been involved in the process of forming synthetic households. Once a household is built, transmission from infectious to susceptible household members is attempted on a daily basis with a probability corresponding to 1 − exp(− *βf*_hh_Δ*t*), where Δ*t* is set to 1 day, 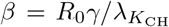, and *f*_hh_ is an enhancing factor that takes into account increased transmission due to school closures and home working. Household members who quarantine or are hospitalized are removed from the transmission chain to free up computational resources, while the rest of the household structure is maintained for future infection events. If during the course of the simulation the search for members of new families runs out of susceptibles, the simulation program retries searching for new members up to a limited number of attempts. Beyond that value, we assume that remaining household members must be recovered individuals and the effective household size is reduced. The maximum number of attempts is limited to 20 for efficiency.

The use of a household structure alters the force of infection: a household will tend to contain the wider spread of the disease, especially if a quarantine or stay-at-home policy is enabled. Since we are breaking the local full mixing of the population, the epidemic curve will follow a less steep growth rate. Thus to make sure that the model reproduces the correct growth rate given the parameters estimated using well-mixed models, a multiplicative term to the force of infection is introduced that bridges between the two descriptions. The term, calculated using a semi-analitical approach and checked with simulations, is discussed in detail in the Supplementary Material SM-1.

### 2.3 Human mobility

We estimate the movement of individuals within each country using the Radiation Model [24, 25] fitted on commuting data. Individual have a set of preferred locations, with an associated frequency of visit, which is assigned at the start of the simulation based on the fluxes between grid elements estimated by the radiation model [26, 27]. Each day a destination is chosen for each individual among his set of preferred locations according to their frequency of visit, and the individual is moved to that preferred location where it participates to the dynamics of that cell for a duration corresponding to the average work day. The individual is then moved back to his original location.

Data on human mobility is available for all countries investigated, through their respective National Statistical Institutions, in the form of commuting matrices between national political subdivisions. This is typically in form of estimates of the number of individiduals who reside in one subdivision and work in another subdivision. For Italy, information on place of study (for 16+ years old) is also available. This information is thus work oriented, however it is worth noting that even in this case it is still an approximation to the true fluxes, since it misses mobility of individuals outside of the typical work shift. Despite these caveats these dataset may be used effectively for the description of the spatial spread of infectious diseases at large scales [28].

Our simulation model uses grid elements as basic areal units, thus we identified the grid cells corresponding to the different national subdivisions specified by the commuting matrices, and calculated the corresponding aggregated fluxes. These fluxes depend on the fraction of the population participating in the global commuting between regions which is a model parameter [26, 27]: the optimal value was identified by maximizing the common part of commuters – a measure of similarity of flow estimates based on the Sørensen index [29, 30, 31]. Further details are available in the Supplementary Material SM-1.

### 2.4 Ascertainment rate and risks of hospitalization and death

We derive the risk for a detected case to be hospitalized as a severe or critical case from age stratified data of Spanish surveillance [32]. We further derive age stratified death risks for severe and critical cases based on both Spanish and Italian data [32, 33]. Finally, we use the Spanish serological survey to estimate ascertainment rates (the percentage of detected cases). By appropriately combining risk percentages with the ascertainment rate, we derive the risk of death for an infected individual, or Infection Fatality Ratio (IFR). Table 1 shows the resulting rates, while all calculations required are detailed in the Supplementary Material SM-2.

**Table 1:** Estimats of ascertainment rate, risk of being severe or critical for detected cases, risk of death for critical or severe cases, and Infection Fatality Ratio (95% confidence interval in brackets). In Grasselli *et al*. [33], which we use to estimate death risks, individuals up to 39 years old are grouped into two groups corresponding to 0-19 and 20-39. No deaths were registered in the youngest group: rates for the 20-29 and 30-39 were obtained from the second group. The Infection Fatality Rate is obtained by appropriately combining the the other five columns.

| Age group | Ascertainment rate | Risk of being a severe case | Risk of being a critical case |
| --- | --- | --- | --- |
| 0-9 | 0.68% [0.47-1.0] | 27.4% [24.5-30.3] | 4.5% [3.1-5.8] |
| 10-19 | 0.77% [0.61-0.98] | 15.5% [13.8-17.3] | 1.4% [0.86-2.0] |
| 20-29 | 5.5% [4.3-7.0] | 10.3% [9.81-10.8] | 0.66% [0.53-0.80] |
| 30-39 | 8.2% [6.5-10] | 15.6% [15.2-16.1] | 1.21% [1.07-1.36] |
| 40-49 | 7.3% [6.3-8.6] | 22.6% [22.1-23.0] | 2.11% [1.96-2.26] |
| 50-59 | 9.1% [7.7-11] | 29.2% [28.8-29.7] | 3.65% [3.47-3.83] |
| 60-69 | 9.4% [7.8-11] | 42.9% [42.3-43.4] | 7.37% [7.10-7.65] |
| 70-79 | 10.7% [8.7-13] | 57.1% [56.6-57.7] | 6.97% [6.70-7.24] |
| 80+ | 29% [21-41] | 44.5% [44.1-44.9] | 0.696% [0.627-0.764] |

Table 1: Estimats of ascertainment rate, risk of being severe or critical for detected cases, risk of death for critical or severe cases, and Infection Fatality Ratio (95% confidence interval in brackets).
| Age group | Risk of death for severe cases | Risk of death for critical cases | Infection Fatality ratio |
| --- | --- | --- | --- |
| 0-9 | 1.2% [0.00-2.9] | 0% [-] | 0.0022% [0.0015-0.0033] |
| 10-19 | 2.8% [1.2-5.1] | 0% [-] | 0.0034% [0.0027-0.0043] |
| 20-29 | 1.8% [1.2-2.5] | 8.9% [3.4-15] | 0.014% [0.011-0.017] |
| 30-39 | 1.9% [1.4-2.4] | 8.9% [5.8-12] | 0.033% [0.026-0.041] |
| 40-49 | 2.47% [2.13-2.81] | 13% [10-15] | 0.061% [0.052-0.071] |
| 50-59 | 5.00% [4.62-5.39] | 16% [15-18] | 0.19% [0.16-0.22] |
| 60-69 | 11.1% [10.6-11.6] | 31.9% [30.1-33.7] | 0.67% [0.56-0.79] |
| 70-79 | 30.7% [30.0-31.4] | 43.6% [41.5-45.6] | 2.2% [1.8-2.7] |
| 80+ | 68.5% [67.9-69.0] | 64% [59-69] | 8.9% [6.4-13] |

### 2.5 Interventions

A number of interventions are implemented following approaches used by various governments to control the disease as well as information available from Google COVID-19 Community Mobility Reports [19]: here we detail the interventions available (contact tracing, social distancing, school closures, stay-at-home including work from home, travel restrictions), while details of the interventions implemented for each country are discussed in the Supplementary Material SM-1. All interventions may be implemented immediately at the time the interventions are activated, or with a linear increase from the time of activation during a specified number of days.

#### 2.5.1 Contact tracing

A rudimentary contact tracing is implemented by recording contacts of each individual being infected and entering the latent class. As the individual progresses to removal, a fraction of his contacts are isolated and quarantined. We assume a delay of about 3 days from the time of case detection (removal) to quarantine of his contacts. Contact tracing is used mostly to simulate the controlled epidemic during the initial phase of the outbreak.

#### 2.5.2 Social distancing and school closures

Social distancing consists in reducing physical contact between people with the aim of reducing transmission. We implement it as a generalized contact reduction of *K*_other_. Closure of schools is implemented by reducing the transmission in schools, typically to zero. In this model schools are not explicitly described, thus reduction of transmission is implemented by multiplying the school contact matrix *K*_school_ by a reduction factor.

#### 2.5.3 Home working and stay-at-home

We consider home confinement for both working and non-working population (the stay-at-home idea). Working individuals may be asked to stay at home and limit their main interaction with member of their household. Similarly, non-working individuals may be asked to remain home. Both confinement measures are implemented mechanistically: we assume that a fraction *f*_*w*_ of working individuals will stay at home and not commute or interact for work, having work transmission reduced to zero. Similarly, individuals who are staying at home due to social distancing, will cease to interact socially and will only have contacts within their household.

#### 2.5.4 Household transmission

The model uses synthetic households as discussed previously. We assume that due to school closures, home working and stay-at-home, household transmission may increase: this is implemented by tuning the multiplicative factor *f*_hh_ introduced earlier.

#### 2.5.5 Travel restrictions

Travel restriction may be implemented by reducing the probabilities *p*_*ij*_ underlying the radiation model (Ref. [24] and Supplementary Material SM-1). Note also that individuals who do not go to work or are quarantined do not move. The application of certain restrictions (for instance home working and stay-at-home) will indirectly contribute to travel reduction. Travel restriction may also be implemented by preventing individuals from moving between certain areas of the country, typically between administrative subdivisions. Travel restrictions are country specific and typically require specific implementations.

#### 2.6 Fitting methods and epidemiological data

We use an approach based on recent work on Gaussian optimization detailed in Refs. [34] and [35], and adapted to our specific needs. The method is particularly advantageous as it reduces the computational time required for the fitting of a factor between 20 and 50, making highly detailed modelling available to fast forecasting. We also use a Sequential Monte Carlo [36] algorithm for verification of the goodness of the fits. Both methods are detailed in the Supplementary Material SM-1. We fit the basic reproductive number *R*_0_, the recovery rate *γ*, and the initial condition by setting an importation rate modulated by province (for Spain and Italy) and Local Authority (for the United Kingdom) and the time span during which importations occur (see Supplementary Material SM-1). Since rates of case confirmation in Italy, Spain and UK may differ, we fit on daily national death counts.

We fit age-dependent susceptibilities using an approximated approach based on the eigenvector corresponding to the maximum eigenvalue of the contact matrix. Specifically, we build a pseudo contact matrix which reproduces the age distribution of cases obtained from a simulations with unitary susceptibilities, and then we fit a set of susceptibilities that alters the age distribution to reproduce the one from data. The method is detailed in the Supplementary Material SM-1.

## 3 Results

The analysis of the data from Spain and Italy allows to estimate ascertainment rate of cases, rates of severe and critical cases and rates of death, to finally determine the underlying infection fatality ratio (Table 1). The latter is the rate of fatality per infected individual, regardless of it being detected or not. A corresponding infection-fatality rate for reported cases can be estimated by dividing the IFR by the ascertainment rate. In the Supplementary material SM-2 we show that the IFR obtained can reproduce observed age distribution of deaths in European countries within reasonable accuracy. In addition to these rates, we also estimated age-dependent susceptibilities for each of the countries discussed: the results are consistent among countries, and allow us to derive average susceptibilities to the SARS-CoV-2 virus usable as reference in future works (Table 2). The results show that susceptibility to the SARS-CoV-2 virus is low for individuals up to 50 years of age, and then climbs rapidly with age.

**Table 2:**
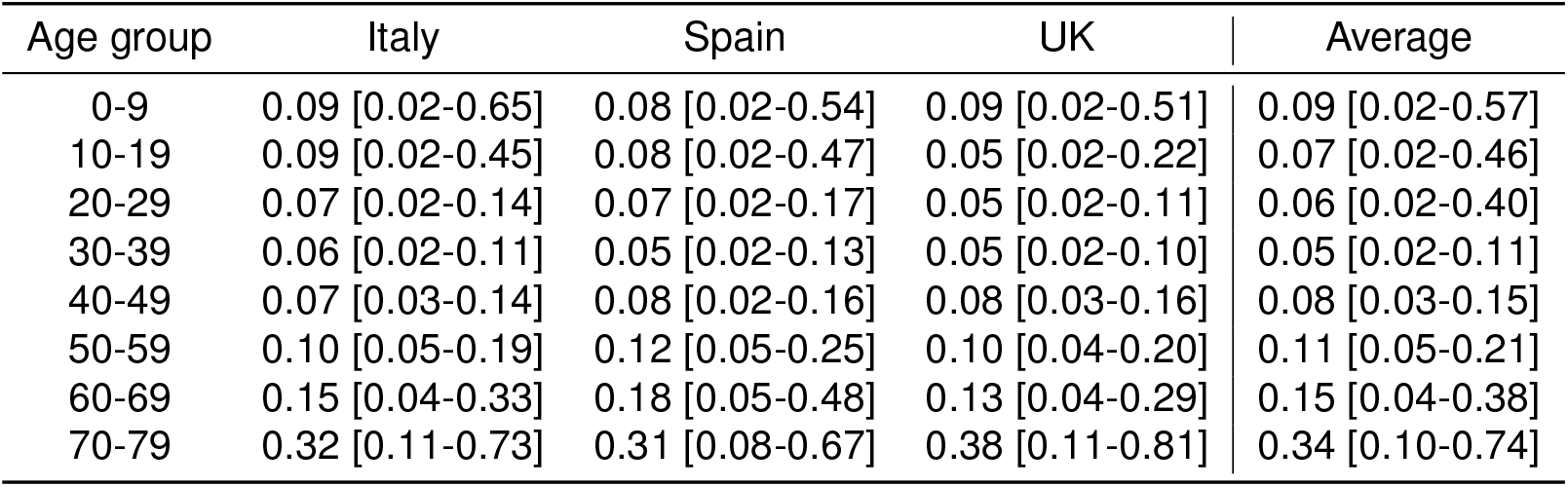
Estimated relative susceptibilities for Italy, Spain and the United Kingdom with respect to the 80+ age group. The last column is an average of the previous three that can be used as reference in future works. Values in brackets represent the 95% confidence interval.

We estimted, for each country, the unrestricted *R*_0_ appearing in the force of infection (1), which would be the *R*_0_ of the disease measured in China in the absence of any containment measure. This represents a reference estimate that is independent of the specific contact pattern of the country under study. We find that the three countries lead to similar estimates of *R*_0_ (Italy: 4.1, 95% CI [2.6-5.6]; Spain 4.3, 95% CI [3.1-5.5]; UK 3.9, 95% CI [3.1-4.9]), while the number of predicted cases is in line with what observed in data. The results for the spatial distribution of cases show a remarkable similarity with data, provided the description of human mobility is adequate. In Italy, the density of cases remains high in the Northern part of the country, especially in the Lumbardy, Veneto and Emilia-Romagna regions which were the areas of Italy that registered the highest number of cases during the peak of the epidemic. In simulations, clusters of cases are also observed in Rome, Naples and other densely populated areas of the south, however the fraction of infected population of these areas remains low. The difference may reflect a fundamental difference in the geographic distribution of the population between the highest infected areas and the rest of the country: the affected areas in the north of Italy form a continuous densely inhabited hinterland, with population densities among the highest in Europe (Lumbardy alone counts one sixth of the population of Italy). This extended area of interconnected highly population densities, coupled with measures that curbed mobility within the country, may have favoured the local spread of the disease whilst containing it within the affected regions.

Results for the United Kingdom also show a remarkable resemblance to the observed spatial cumulative incidence (Fig. 5). The initial stages of the epidemic are dominant in the highly populated areas of England, and spread towards lower populated areas during the declining phase following restrictions. The United Kingdom was subject to several importations from abroad (evidence of this is also suggested by a recent report on genomic studies [37]) that led to a widespread distribution of cases. The United Kingdom presents a high population density in and around large centres in England, with lower population densities in Wales and Scotland; restriction in the United Kingdom were less severe than those imposed in Spain and Italy, and mobility level were higher than those observed in the other two countries, especially towards the end of the lockdown period: in simulations, this is reflected in the spread towards low populated areas during the later stages of the declining phase.

**Figure 4:**
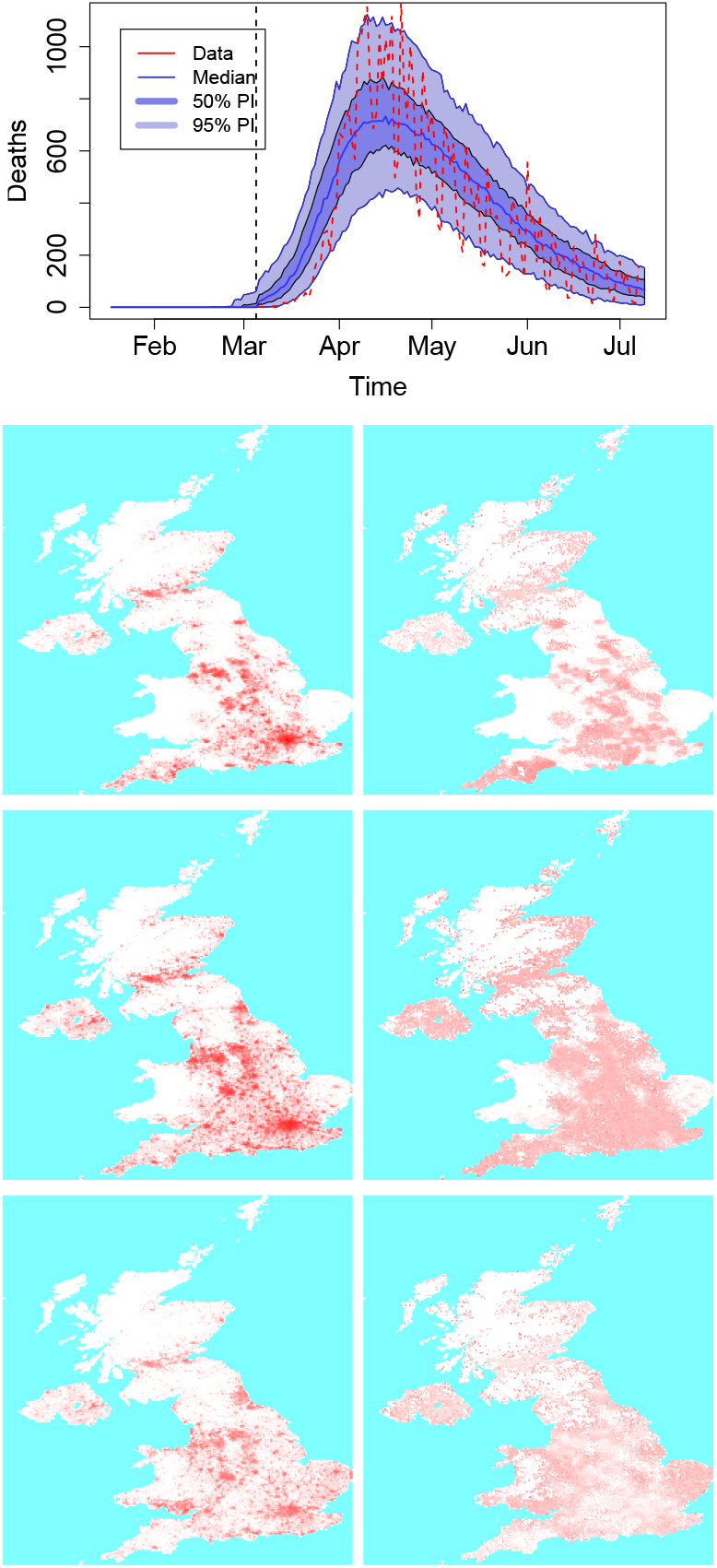
(Top) Country level dynamics of SARS-COV-2 inferred by death incidence data for the UK. Fitted parameters are *R*_0_ = 3.9 (95% CI [3.1 - 4.9]), *t*_0_ = 14 (95% CI [2 - 24]) days, *τ*_*γ*_ = 3.9 (95% CI [1.3-7.5]), and log *ω* = *−*5.2 (95% CI [−7.6 – −2.8]) days^−1^. (Bottom) Frames from an average of 10 simulation for the UK, corresponding to cases at the end of week 3, 7 and 12: on the left is the average incidence, on the right the incidence as a fraction of the population.

**Figure 5:**
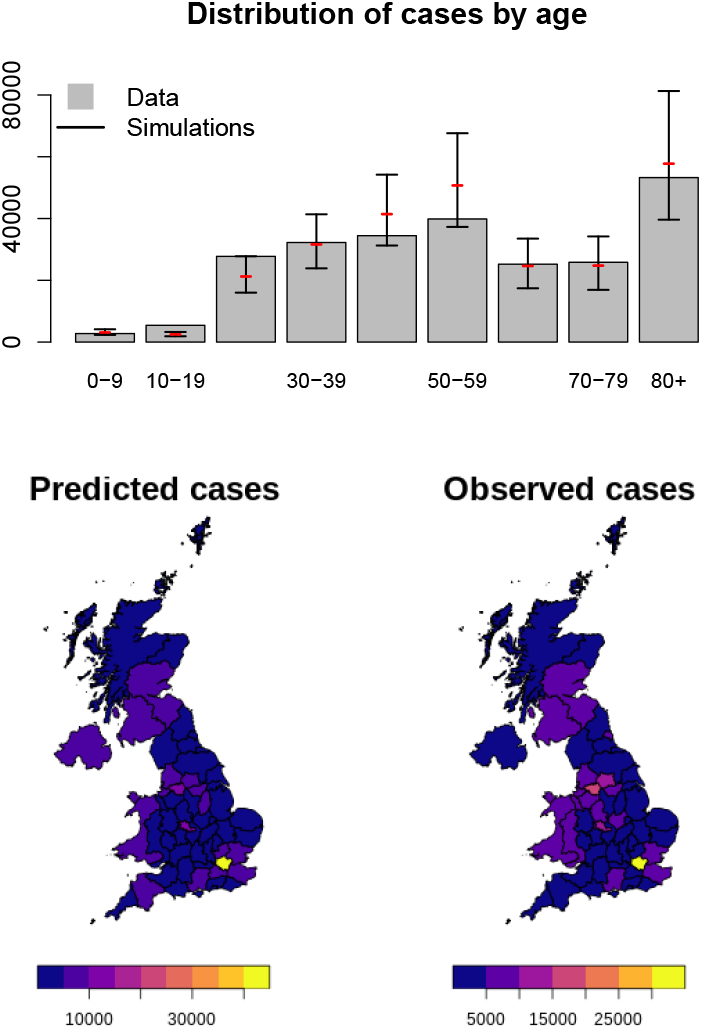
(Top) Age distribution of cases predicted by the model and comparison with data from the United Kingdom. The model overestimates cases in the middle age groups, and underestimates cases in the elderly. (Bottom) Comparison between predicted spatial spread of COVID-19 cases and observed distribution.

**Figure 6:**
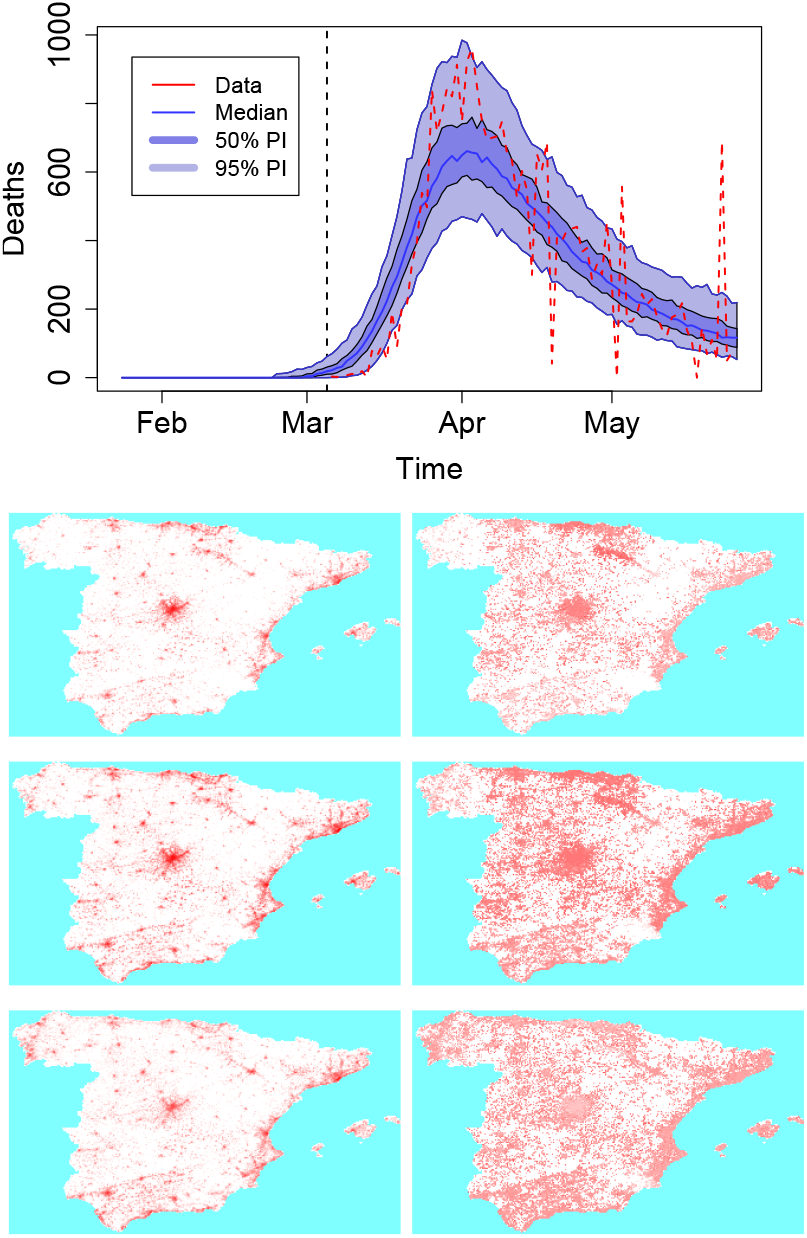
(Top) Country level dynamics of SARS-COV-2 inferred by death incidence data for Spain. Fitted parameters are *R*_0_ = 4.3 (95% CI [3.1 - 5.5]), *t*_0_ = 13 (95% CI [1 - 17]) days, *τ*_*γ*_ = 3.9 (95% CI [0.4-7.6]), and log *ω* = *−*7.4 (95% CI [−9.3 – −5.7]) days^−1^. (Bottom) Frames from an average of 10 simulations for Spain, corresponding to cases at the end of week 2, 4 and 7: on the left is the average incidence, on the right the incidence as a fraction of the population.

**Figure 7:**
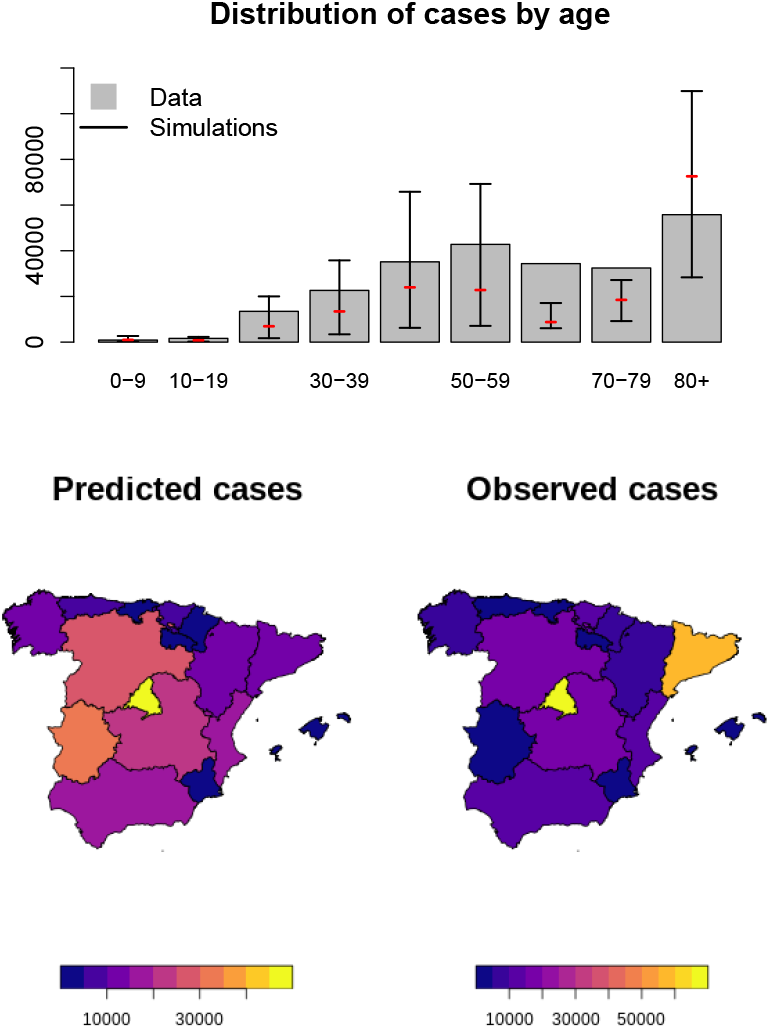
(Top) Age distribution of cases predicted by the model and comparison with data from Spain. The model overestimates cases in the middle age groups, and underestimates cases in the elderly. (Bottom) Comparison between predicted spatial spread of COVID-19 cases and observed distribution.

Simulation for Spain fail to reproduce the correct spatial spread of the SARS-COV-2: this is likely a direct consequence of the inadequacy of the radiation model to correctly describe mobility within the country. Direct inspection of commuting data for Spain reveals that most commuting occurs between a few specific provinces (for instance between Madrid and a few of its neighbouring provinces), whilst the rest of the country is subjected to a substantially lower level of transmission. This makes it hard to any model based purely on distance and population size, like the gravity model, the radiation model and their descendants, to provide a correct description of mobility that is valid countrywide. In our simulations, the highest populated area of Madrid has the highest count of cases, however rural and less densely inhabited areas of the country are subject to diffused local transmission. Similarly to the United Kingdom, importation of initial cases was more widespread in the country with respect to Italy, with cases occurring in provinces with low density population. Mobility of individual greatly diminished in the middle of March but increased again during the following weeks: spatial spread on short scales can be observed during the declining phase of the epidemic.

## 4 Discussion

Our estimates of the IFR obtained from Spanish and Italian data differ from the early estimates of Verity *et al*. [16]: the rates found suggest a lower IFR for middle aged individuals (30-60) and a higher IFR for 80+ individual with respect to the Verity estimates. Our estimates take advantage of the Spanish serological survey, and thus do not require to estimate the fraction of asymptomatic infections that occur within a population – a fraction that still today is not well quantified. Nevertheless, our IFR reproduce reasonably well the age distribution of deaths registered in several countries. We estimated also an age-dependent susceptibility that shows a marked dependence on age. The estimated susceptibilities confirm that young individuls are less susceptible than older individuals to the virus, and they further suggest that this low susceptibility is shared by middle aged individuals (30-60 years old) that instead account for a large fraction of cases. The reason of this apparent contradiction is acutally due to the fact that in our modelling approach we distinguish between working and non-working individuals, with the first subject to more mixing due to differential measures imposed by government.

Previous studies have reported a range of susceptibilities as discussed in the review by Russell *et al*. [38]. The authors of the review report an odd of being an infected contact in young individuals less than 19 year old compared to adults of age greater than 20 years old of 0.56 (95 % CI 0.37 - 0.85), pooled from all reported studies. Based on our susceptibilities, one can build the average susceptibility Θ_*Y*_ of age set *Y*. which is simply an average over the population sizes Θ_*Y*_ = ∑_*j*ϵ*Y*_ *N*_*j*_*σ*_*j*_*/*∑_*j*ϵ*Y*_ *N*_*j*_, where *N*_*j*_ is the size of age group *j*. The ratio Θ_≤19 y.o._*/*Θ_≥20 y.o._ is the relative odds ratio of being an infected contact between children and adults: focussing for simplicity on Spain, one gets a value of *≈* 0.43 which is not far from the value found by Russell *et al*. [38].

The model uses an agent based description of the country population that incorporates households, as well as interventions adopted by the various government, or changes in contact patterns measured by social data. Some of the parameters we use are based on previous estimates: for instance the age-mixing matrices are based on the work of Prem *et al*. [22] which are projections from socio-economical data. Reduction in contact patterns are based on estimates from social data which we interpret as a direct measure of such reduction. The simulation model includes importations from other countries and takes into account, to some extent, the reduction of air travel between countries: the spatial distribution of imported cases reflects the distribution of the initial cases observed in each country. Super spreading events are not explicitly included in this approach, but their effects are implicitly incorporated by the modulated importations in the initial stages of the simulation.

We adapt modern methods for Bayesian optimization that provide fast estimates of model parameters, permitting to simulate and estimate parameters for a country of roughly 60 million individuals using detailed agent based simulations in a relative short simulation time. Despite the simulations being parallelized, the version of the main fitting algorithm implemented, based on Gaussian processes, is currently not parallelized: methods for parallelizing Gaussian processes do exist. The simplest option would be to draw multiple points from the current posterior variance and progressively update it as the evaluation of each point completes. This would allow even faster results despite the model complexity.

Our simulation model is capable of reproducing several features of the spread of COVID-19 in three distinct European countries: the spatial spread, the temporal dynamics of the epidemic, the age distribution of cases. We estimated an unrestricted and country independent basic reproductive number *R*_0_ (referenced to China) which is higher than previous estimates in China as well as other European countries. This might be linked to the fact that the model simulates the dynamics of both detected cases and undected cases, whilst estimates have mostly used the available data on reported cases. According to the IFR that we obtained from the Spanish and Italian data, this under-represents the spread of the virus in an age-depentent fashion, which might lead to a different *R*_0_. It is worth noting that recent research has led to estimates of *R*_0_ higher than the early reported ones [39, 40].

The spatial spread of the disease is correctly described as long as mobility can be described by an underlying model for human mobility. The latter was modelled using the radiation model, preferred to the gravity model due to the need of aggregating fluxes between grid elements up to the level of counties. For Italy and the United Kingdom, the radiation model achieves a CPC of about 0.45 which, considering the use of an aggregation process from base areal units, is in line with the performance of both the radiation and gravity model. However, the limitations of the radiation model are evident in the description of mobility in Spain that only scores a CPC of 0.21. Possibly, the use of more recent models for human mobility might mitigate such discrepancies [30].

The agreement of the *R*_0_ obtained from the independent fits performed in each country as well as the capability of the model of reproducing the observed data suggest its possible use in other contexts. One possibility could be to estimate the impact of future interventions in the countries studied in this work or other European countries, provided age-stratified information on cases and deaths from COVID-19 as well as information on the mobility of individuals are available for such countries. Another possibility could be its use in Low and Middle income countries: the key ingredients that would permit this are the availability of contact matrices from the work of Prem *et al*. [22] for almost all countries worldwide, the assumption that age-dependent susceptibility does not depend on anything other than age, and similarly the assumption that the IFR of COVID-19 derived from the Spanish data could be exported to other countries. Under these conditions the model could be used to explore the spread of COVID-19 in African and South American countries. Data on human mobility should also be available to permit studies of spatial spread, but since this is often not readily available in these contexts, with a bit of effort one could use spatially detailed time series of detected cases to infer the level of human travel by fitting an appropriate model for mobility.

## Data Availability

All data is publicly available as detailed in the references. The code is available at the following repository: https://github.com/andreaparisi-science/SpatialCOVID19

## Acknowledgements

The authors wish to thank Erin Gorsich, Trystan Leng, Hector McKimm, Bridget Penman and Emma Southall for sharing their reviews on recent literature of COVID-19. The work was funded by the National Institute for Health Research (NIHR) (project reference 17/63/82) using UK aid from the UK Government to support global health research. The views expressed in this publication are those of the authors and not necessarily those of any of the funders.

