## Supplementary Material SM1 for "Spatially resolved simulations of the spread of COVID-19 in European countries"

#### Supplementary Material S1

### A Methods

#### A.1 Epidemiological model

The disease is described using an age structured compartmental model comprising 11 distinct categories and 9 age groups. Each age group represents a ten year interval (0-9, 10-19, etc), with the last representing the population over 80 years old. The choice is dictated by the stratification used for data on hospitalized cases and deaths [1, 2, 3]. The epidemiological model is schematically described in the main text. Susceptible individuals (S) become exposed (E) upon transmission of disease from an infected individual. Exposed individual remain in the exposed class for $1/\sigma$ days after which they transition to either a detected infection (I) with probability $z_i$ or an undetected infection (U) with probability $1 - z_i$, where $z_i$ is the age-dependent ascertainment rate. Both detected and undetected cases may transmit the disease to susceptible individuals with transmission rate $\beta = R_0\gamma/\lambda_{\rm CH}$ and susceptibility $\sigma_i$ as explained in the main text. All cases remain infectious for an average of $1/\gamma$ days, after which they are removed from the transmission chain. Undetected cases move to an undetected-removed class (Z), while detected cases may quarantine at home (Q), become serious (H) or critical cases (C). Those who become serious or critical, do so with a delay which is taken into account with a waiting class (W). The probability of a detected case to home quarantine is $q_i$, and that of a non-quarantined detected case to become a serious case is $h_i$. These two probability are calculated from the probabilities of a detected case to become a hospital case $\hat{h}_i$ or a critical case $\hat{c}_i$ as

$$
\begin{aligned}
q_i &= 1 - \hat{h}_i - \hat{c}_i \\
h_i &= \hat{h}_i/(\hat{h}_i + \hat{c}_i)
\end{aligned}
$$

The average permanence in the waiting class W is $1/\rho$. The average permanence in hospital and critical care is respectively $1/\mu_h$ and $1/\mu_c$. Age dependent death probabilities for individuals in hospital and critical care are respectively $d_j^h$ and $d_j^c$. Transition between exposed to infectious classes and from infectious classes to subsequent classes may be implemented using multiple classes to provide gamma distributed transitions, although we did not take advantage of this. The full model is described in figure 1.

| | | | | |
|---|---|---|---|---|
| $S_i$ | $\xrightarrow{R_0^*\gamma K_{ij}(I_j^q+A_j^q)/\lambda_{K_{\mathrm{CH}}}N_j}$ | $E_i^1$ | | Non-work transmission |
| $S_i$ | $\xrightarrow{R_0^*\gamma K_{ij}^{\mathrm{work}}(I_j^q+A_j^q)/\lambda_{K_{\mathrm{CH}}}N_j}$ | $E_i^1$ | | Work transmission |
| $S_i$ | $\xrightarrow{R_0^*\gamma f_{\mathrm{fam}}K_{ij}^{\mathrm{home}}(I_j^q+A_j^q)/\lambda_{K_{\mathrm{CH}}}N_j}$ | $E_i^1$ | | Household transmission |
| $E_i^q$ | $\xrightarrow{L\sigma}$ | $E_i^{q+1}$ | $1 \leq q < L$ | Multiple Exposed classes |
| $E_i^L$ | $\xrightarrow{L\sigma z_i}$ | $I_i^1$ | | Transition to case |
| $E_i^L$ | $\xrightarrow{L\sigma(1-z_i)}$ | $U_i^1$ | | Transition to undetected |
| $I_i^q$ | $\xrightarrow{L\gamma}$ | $I_i^{q+1}$ | $1 \leq q < L$ | Multiple case classes |
| $U_i^q$ | $\xrightarrow{L\gamma}$ | $U_i^{q+1}$ | $1 \leq q < L$ | Multiple undetected classes |
| $I_i^L$ | $\xrightarrow{h\,L\gamma}$ | $W_i$ | | Transition to severe or critical case |
| $I_i^L$ | $\xrightarrow{(1-h-c)\,L\gamma}$ | $Q_i$ | | Quarantining |
| $W_i$ | $\xrightarrow{h\,L\gamma}$ | $H_i$ | | Hospitalisation |
| $W_i$ | $\xrightarrow{c\,L\gamma}$ | $C_i$ | | Entering critical care |
| $U_i^L$ | $\xrightarrow{L\gamma}$ | $Z_i$ | | Removal of undetected |
| $H_i$ | $\xrightarrow{d_h\,\mu}$ | $D_i$ | | Death of severe case |
| $H_i$ | $\xrightarrow{(1-d_h)\,\mu}$ | $R_i$ | | Recovery of severe case |
| $C_i$ | $\xrightarrow{d_c\,\mu}$ | $D_i$ | | Death of critical case |
| $C_i$ | $\xrightarrow{(1-d_c)\,\mu}$ | $R_i$ | | Recovery of critical case |
| $Q_i$ | $\xrightarrow{\zeta}$ | $R_i$ | | Recovery of quarantined |

Figure 1: Full set of stochastic transitions implemented. There are three sets of transitions involving transmission: the first two are implemented separately because each has a distinct associated action: the work transmission for instance only occurs if individual are not staying at home, whereas non-work transmission may still occur for all individuals, depending on the set-up. Household transmission is implemented separately from the other two. The waiting class W describes the delay to hospitalization.

#### A.2 Household attack rate and modifications to the force of infection

The use of a household structure alters the force of infection: a household will tend to contain the wider spread of the disease, especially if a quarantine or stay-at-home policy is enabled. Since we are breaking the local full mixing of the population, the epidemic curve will follow a less steep growth rate. Thus to compare the household model with the well mixed description we need to introduce an effective basic reproductive number that reproduces the same growth rate of the well mixed model: this allows to parametrize the model with the estimates obtained using simpler well mixed models.

The basic reproductive number $R_0^{\mathrm{eff}}$ is the maximum eigenvalue of the matrix

$$\begin{aligned} Z_{ij}(\alpha) &= \frac{R_0^{\mathrm{CH}}}{\lambda_{\mathrm{K}}}\left(K_{ij}^{\mathrm{work}} + K_{ij}^{\mathrm{school}} + K_{ij}^{\mathrm{other}} + \alpha K_{ij}^{\mathrm{home}}\right) \\ &= \frac{R_0^{\mathrm{CH}}}{\lambda_{\mathrm{K}}} K_{ij}(\alpha) \end{aligned} \tag{1}$$

for an appropriate value of $\alpha$, where $R_0^{\mathrm{CH}}$ is the estimated pre-lockdown basic reproductive number for China, and $\lambda_{\mathrm{K}}$ is the maximum eigenvalue of the full Chinese contact matrix. It follows that $R_0$ evaluated for the well mixed model in the country of interest is the maximum eigenvalue of the matrix $Z_{ij}(\alpha)|_{\alpha=1}$. On the contrary, when $\alpha = 0$ the model has no transmission in households. Thus the correct value when including a household description is in between these two extremes. The factor $\alpha$, representing the contribution of $K_{ij}^{\mathrm{home}}$ to the transmission, is related to the fraction of individuals who will become infected within a household. Given a household of mean size $\mu$, if the transmission probability per individual per day were 1, then at most there would be $\mu - 1$ secondary infections. Thus, the contribution of $K_{ij}^{\mathrm{home}}$ would be multiplicatively bounded by $\alpha = (\mu - 1)\gamma/\lambda_{K^{\mathrm{home}}}$. Note that such ratio must be necessarily less than one.

Suppose now that the probability of transmission per individual per day is less than 1, then the average number of secondary infections within a household will be $\mu^* < \mu - 1$. We can evaluate $\mu^*$ numerically by randomly sampling individuals from the age structured population.

| Parameter | Use | Value |
|---|---|---|
| $R_0$ | Basic reproductive number | fitted |
| $1/\sigma$ | Permanence in the exposed class | 3 days |
| $1/\gamma$ | Average duration of transmission window | fitted |
| $1/\rho$ | Delay to hospitalization | 3 days |
| $z_i$ | Ascertainment rate | 0.68% - 29% |
| $\hat{h}_i$ | Probability to be a severe case if detected | 10.3% - 57.1% |
| $\hat{c}_i$ | Probability to be a critical case if detected | 0.66% - 7.37% |
| $1/\zeta$ | Average time to removal if undetected | 2 days |
| $1/\mu_h$ | Average permanence in hospital ward | 6 days |
| $1/\mu_c$ | Average permanence in critical care | 7 days |
| $d_i^h$ | Probability of dying if hospitalised | 1.2% - 68.5% |
| $d_i^c$ | Probability of dying if critical | 0.0% - 64% |

Table 1: List of parameters and values used in simulations. When a range of value is provided, this is due to the fact that the parameter is age dependent.

| Country | Mean size of household | Household attack rate | $R_0^{\rm eff}/R_0$ [95% C.I.] | $\beta^{*,\rm HH}/\beta^*$ |
|---|---|---|---|---|
| Italy | 2.31 | 0.1948±0.0005 | 1.23730 [1.23728–1.23732] | 1 (a) |
| Spain | 2.5 | 0.1976±0.0005 | 1.26042 [1.26040–1.26045] | 1 (a) |
| UK | 2.4 | 0.2020±0.0005 | 1.28792 [1.28789–1.28795] | 1 (a) |
| Wuhan | 2.85 | 0.1968±0.0005 | 1.25459 [1.25456–1.25461] | 1 (a) |
| Italy | 2.31 | 0.3000±0.0006 (b) | 1.23311 [1.23308–1.23313 ] | 1.639±0.003 |
| Spain | 2.5 | 0.2997±0.0006 (b) | 1.25531 [1.25528–1.25534] | 1.609±0.003 |
| UK | 2.5 | 0.3002±0.0006 (b) | 1.28205 [1.28202–1.28209] | 1.575±0.004 |
| Wuhan | 2.85 | 0.3000±0.0006 (b) | 1.24933 [1.24931–1.24936] | 1.615±0.003 |

Table 2: Corrective coefficient to the general and household transmission rates, based on average household size and country. (a) For the first set of values, the household transmission rate was imposed to be the same as the general transmission rate. (b) The second set of values has the attack rate set to converge to 0.30.

We build households around infected individuals by choosing the household size according to a Poisson distribution with mean equal to the mean household size, and other susceptible members according to the age distribution $g(i) = K_{ij}^{\rm home}/\sum_i K_{ij}^{\rm home}$, where $j$ is the age group of the infected individual. Since household members are chosen according to $g(i)$, the distribution of contacts within households reflects the $K_{ij}^{\rm home}$ matrix. For each household we attempt, each day, transmission from the primary infected to other household members at rate $\beta$, during a period of $1/\gamma$ days. Finally we count the final fraction of infected in each household excluding the infector: this represents the household attack rate $\nu$. The attack rate can be used to evaluate $\alpha$ via

$$\alpha = \nu\mu^*\gamma/\lambda_{K^{\rm home}} \tag{2}$$

By repeating the procedure 1,000 times using each time 1,000 households, we get an initial estimate for the household attack rate. Using this value we get the ratio between $R_0$ and the effective $R_0^{\rm eff}$ – the effective reproductive number associated with the household description.

$$\frac{R_0}{R_0^{\rm eff}} = \frac{\max_{\lambda\in\sigma(Z_{ij}(\alpha)|_{\alpha=1})}\lambda}{\max_{\lambda\in\sigma(Z_{ij}(\alpha)|_{\alpha^*})}\lambda} \tag{3}$$

This ratio can also be estimated by directly simulating early growth with a fully well mixed transmission model, leading to values comparable to the ones reported in table 2. Since the reproductive number is proportional to the transmission rate, to correct the growth rate of the early epidemic so that it reflects the growth rate of the corresponding well mixed model for a given $R_0$, it is sufficient to increase the per contact transmission rate to $\beta^* = \beta R_0/R_0^{\rm eff}$, so that the new effective reproductive number $R_0^{*,\rm eff}$ becomes equal to $R_0$. The household attack rate $\nu^*$ resulting from the new per contact transmission rate can be readily evaluated (Table 2).

The value for the household attack rates are higher than that measured by Bi *et al.* . [4] (11.2% 95% C.I. [9.1-13.8]), but lower than the 30% estimated in Ref. [5]. The R script that was used to evaluate the above results, can also force the household attack rate to a specific value, if this is available, by further tuning the within household transmission rate $\beta^{*,\rm HH}$. Since the reproductive number is not a linear function of the household attack rate (due to the small finite size of households), the correct values for $\beta^*$ and $\beta^{*,\rm HH}$ must be found iteratively.

We introduce a sequence of household transmission rates $\beta_t^*$, $\beta^{*,\rm HH}$, attack rates $\nu_t^*$ and corresponding effective reproductive numbers $(R_0^{\rm eff})_t$: we set $\beta_0^* = \beta$ and $\beta_t^* = \beta_{t-1}^* R_0/(R_0^{\rm eff})_t$.

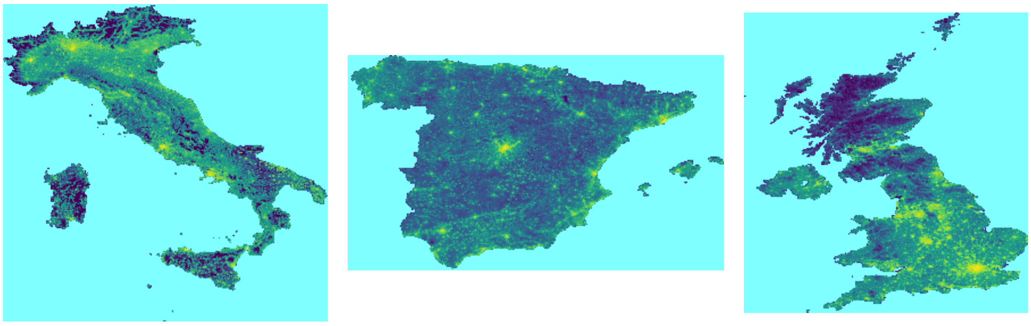

Figure 2: Population distribution for Italy, Spain and the United Kingdom. The three countries are represented under the same spatial scaling using an equirectangular projection. The spatial resolution is 5km.

We further set $\beta_t^{*,\mathrm{HH}}/\beta_t^* = (\beta_{t-1}^{*,\mathrm{HH}}/\beta_{t-1}^*)\,\hat{\nu}/\nu_{t-1}^*$, where $\hat{\nu}$ is the target attack rate. The sequences $\beta_t^*$, $\beta^{*,\mathrm{HH}}$, $\nu_t^*$ and $(R_0^{\mathrm{eff}})_t$ converge rapidly to their limit values. Using this method we evaluated how much the within household transmission rate $\beta^{*,\mathrm{HH}}$ should be increased with respect to the general transmission rate $\beta^*$ for an attack rate of 0.30. As shown in table 2, the within household transmission rate is roughly 1.6 times the general transmission rate. This suggests that increased contact from lockdown restrictions might be at the source of such increase. We thus assumed that household transmission increased during lockdown. Taking into account the increase in number of hours of close contact between family members we assumed a peak of 100% increase in contact when both school closures and stay-at-home policies are implemented.

When at home, at the start of the day, each individual will choose one of his preferred locations and will move there for a number of hours corresponding to the average work shift. Individuals who move for more days will stay in the target location for the required number of nights before moving back. While in their target locations individuals will be subject to the local transmission dynamics.

#### A.3 Human Mobility

We use a radiation model [6, 7] for human mobility to describe fluxes of individuals. Our base areal units are the elements of the gridded map (Fig. 2). The probability that an individual in $i$ will travel to grid element $j$, according to the radiation model, is given by:

$$p_{ij} = \frac{N_c}{N} \frac{n_i n_j}{(n_i + s_{ij})(n_i + s_{ij} + n_j)}$$

where $n_i$ is the population at grid element $i$, $s_{ij}$ is the population within a circle of radius $r_{ij}$ centered in $i$, excluding locations $i$ and $j$, and the mobility ratio $N_c/N$ is a fittable parameter.

To reproduce the fluxes, we identified the grid cells corresponding to the different subdivisions, and calculated the resulting aggregated fluxes. These fluxes depend on $N_c/N$: the optimal value was identified by maximizing the Common Part of Commuters – a measure of similarity used in various works on human mobility modelling [8, 9, 10]:

$$CPC(\{T_{ij}\}, \{T_{ij}^*\}) = \frac{2\sum_{ij} \min\left(T_{ij}, T_{ij}^*\right)}{\sum_{ij} T_{ij} + \sum_{ij} T_{ij}^*} \tag{4}$$

where $T_{ij} = n_i p_{ij}$ is the estimated flux between locations $i$ and $j$, and $T^*_{ij}$ is the true flux from data. Assignment of areal units to provinces was achieved by applying a point-in-polygon algorithm for each grid cell with the polygons describing the shape of each province. A proximity algorithm was used to assign any grid cells that was left out because had its centre slightly outside of these polygons. Pre-calculation of bounding boxes for each polygon was used to speed-up the assignment.

For Italy, the 2011 Census provides the number of working individuals commuting daily between provinces [11]. Province boundaries were electronically downloaded from the web site of the Istituto Nazionale di Statistica [12]. For Spain we used microdata from the 'Encuesta de Poblacion Activa', which is a continuous detailed survey of the Spanish population published every three months [13]. The microdata contains information on the provinces of residence and work, from which commuting fluxes can be derived. We built commuting matrices by combining data from the whole 2018 and 2019. Province boundaries are available in electronic format from the Spanish Instituto Geográfico Nacional [14]. Commuting data for the UK is obtained by the home-work commuting matrices available from the 2011 Census [15]. Based on information of residence and place of work of a sample of the population in each Local Authority, these matrices provide an estimate of the number of individuals commuting between the various Local Authorities for work. Polygons for Local Authorities are available through two sets of files [16, 17] that can be electronically downloaded. Differently from other countries, Local Authorities (which represent a basic administrative subdivision in the UK), vary hugely in size, with the City of London extending for just 2.90 km$^2$, and the Highlands in Scotland, extending over 25,657 km$^2$. We thus aggregated commuting data and Local Authorities to a more spatially homogeneous set, corresponding to Ceremonial Counties in England, and NUTS2 level regions in Scotland and Wales.

For all countries in this study, data represents daily trips from home to work. The time spent at destination reflects the average duration of a work day in the each country [18].

#### A.4 Interventions

Interventions reflect both actions taken by government as reported by online news outlets (La Repubblica, Il Corriere, BBC, The Guardian, El Paiś [19]) and behavioural changes as measured by data. Tables 3, 4 and 5 show the set of interventions implemented for Italy, the United Kingdom and Spain. In all cases, school closures were implemented immediately on the day the measures was activated by the government, and household transmission was increased to reflect accordingly increased transmission. Stay at home for working individuals reflected data from the Google COVID-19 Community Mobility Report for workplaces, while social distancing as well as stay at home for non working age groups reflected the Google data for retail and recreation. In Italy and Spain lockdown measures prevented movement between all the 20 Italian regions and all 48 Spanish provinces: the reduction of travel can be estimated to be at least 50-60% in the Veneto region based on data from one major telecom operator [20, 21]. We thus assumed that for both countries the resulting travel reduction was already taken into account by the stay-at-home policy.

#### A.5 Hospitalization, critical care and death rates

The Spanish government made available detailed age stratified data on admissions to hospital and critical care, and overall death rates until to the middle of May [1]. We use the latest data available to obtain a crude estimate of the age stratified rates of critical care admissions for detected COVID-19 cases. Due to the epidemic being far advanced beyond the peak and

| Timeline | Day | Intervention | Delay | Extent | Target |
|---|---|---|---|---|---|
| 23rd Feb | 7 | Tracing probability[a] | 0 | Local | 1.00 |
| | | Travel reduction | 0 | Local | 0.90 |
| | | Household transmission | 0 | Local | 2.00 |
| | | School closures | 0 | Local | 1.00 |
| 24th Feb | 8 | Reduce importations[b] | 12 | National | 1.00 |
| 27th Feb | 11 | Tracing probability[a] | 7 | National | 0.00 |
| 1st Mar | | Household transmission | 0 | Province | 2.00 |
| | | School closures | 0 | Province | 1.00 |
| 4th Mar | 17 | School closures | 0 | National | 1.00 |
| | | Household transmission | 0 | National | 2.00 |
| 23rd Feb | 7 | Social distancing | 0 | Local | 0.80 |
| | | Stay at home work/other | 0 | Local | 0.80 |
| 24th Feb | 8 | Social distancing | 0 | Province | 0.15 |
| | | Stay at home work | 0 | Province | 0.10 |
| | | Stay at home other | 0 | Province | 0.15 |
| 1st Mar | 14 | Social distancing | 14 | National | 0.85 |
| 1st Mar | | Stay at home work | 14 | National | 0.65 |
| 1st Mar | | Stay at home other | 14 | National | 0.85 |
| 4th Mar | 17 | Social distancing | 5 | National | 0.20 |
| | | Stay at home work/young/old | 5 | National | 0.20 |

Table 3: Detail of interventions implemented in simulations for Italy. "Day" measures the number of days from the 16th of February 2020, the day when the first case detected attended hospital. "Delay" is the time interval during which the intervention increases linearly to the "Target" value: a value of zero means the intervention applies immediately. "Extent" refers to the application of the intervention on the Italian territory: "Local" interventions apply to the first 11 municipalties put in lockdown: these are within an area corresponding to two grid elements around Codogno. "Province" refers to the provinces of Lombardia, Veneto and Emilia-Romagna that were declared yellow zones on the 1st of March. "National" interventions are valid over the whole of Italy. Values for social distancing and stay-at-home policies are derived by Google data.

[a] We assume contact tracing was active during the initial stages of the outbreak after the first detected cases. Strict lockdown measures were imposed on the first municipalities. On the 26th of March it was announced that swab would be taken only from symptomatic cases: we assume an interval of about 1 week to bridge with the results from Ref. [22]. Testing of non-symptomatics resumed after the 29th [22], but not because of contact tracing.
[b] We assume a reduction of international travels to Italy, coinciding to travel bans from Italy: we assume that the bulk of transmission afterwards is internal to the country.

due to the typical delay in admission to critical care being just a few days, we assume these crude estimate to be a good approximation of the true estimates. Similarly we get the crude death rates for hospitalized patients irrespective of critical care, and assume them to be a good approximation of the true rates.

To estimate death rates for critical care, we use a study on 1951 patients in Intensive Care Units in Lombardy, Italy [3]. The study provides, age stratified, the number of patients recov-

| Timeline | Day | Intervention | Delay | Extent | Target |
|---|---|---|---|---|---|
| 2nd Mar | -3 | Reduce Importations | 35 | National | 0.00 |
| 12th Mar | 7 | Tracing probability | 0 | National | 0.00 |
| 21st Mar | 16 | School closure | 0 | National | 1.00 |
| | 16 | Household transmission | 0 | National | 2.00 |
| 10th Mar | 5 | Social distancing | 14 | National | 0.77 |
| | 5 | Stay at home work | 14 | National | 0.62 |
| | 5 | Stay at home other | 14 | National | 0.77 |
| 24th Mar | 19 | Stay at home work | 14 | National | 0.69 |
| 7th Apr | 33 | Stay at home work | 14 | National | 0.60 |

Table 4: Detail of interventions implemented in simulations for the United Kingdom. "Day" measures the number of days from the 5th of March 2020, the day when the death occurred. "Delay" is the time interval during which the intervention increases linearly to the "Target" value: a value of zero means the intervention applies immediately. "Extent" refers to the application of the intervention on the UK territory: all interventions for the UK are "National" i.e. they apply to the whole UK territory. Values for social distancing and stay-at-home policies are derived by Google data.

ered, dead and still in intensive care. Admission to intensive care occurred between the 20 of February and the 18th of March (for a total of 28 days), and the study reports on the situation on the 25th or March. To estimate death rates we take into account that the average length of stay in ICU for patients who died was 7 days [3] and that $R_0$ in Lombardy was estimated to decrease from 3.0 around the 20th of February down to 1 in the middle of March [23]. Using a linear approximation for the decrease, the epidemic growth rate may be written as:

$$I(t) = I(0)\ \exp^{(R_0-1)t\left(1-\frac{t}{2\Delta t}\right)\gamma}$$

where $\Delta t$ corresponds to the 28 days interval above. We use this expression to extract critical care admission times for patients in each age group, and we assign a permanence in critical care before death extracted from a Poisson distribution with rate 7 days. We then look at the sum of the two to get all potential deaths before the 25th of March: these would be the number of deaths corresponding to a death rate 1 in each age group. To estimate the age dependent death probability, we tune the fraction of recovered individuals who are expected to die, matching the estimated deaths by the 25th of March in each age group with those observed.

Armed with these death rates one can calculate the estimated number of deaths per age group based on the number of critical cases in Spain, and by subtracting these from the total number of deaths in each age group, one gets the number of deaths of severe (non critical) cases and relative death rates.

A recent serological survey in Spain provides estimates by age group of the fraction of individuals who have been infected by the SARS-CoV-2 virus. The survey gives us an additional key piece of information that we use in these simulations. Combining the data with the number of cases detected one gets the age stratified ascertainment rate of COVID-19. Thus, multiplying the ascertainment rate by the rate of hospitalization (which is relative to detected cases), one get the rate of hospitalization per infected individual, independent on the ascertainment rate. In the Supplementary Material SM-2, we provide the full R notebook that perform the calculations

| Timeline | Day | Intervention | Delay | Extent | Target |
|---|---|---|---|---|---|
| 5th Mar | 0 | Tracing probability | 0 | National | 0.00 |
| 10th Mar | 5 | Reduce importations | 6 | National | 0.14 |
| 12th Mar | 7 | School closure | 0 | National | 1.00 |
| | 7 | Household transmission | 0 | National | 2.00 |
| 16th Mar | 11 | Reduce importations | 7 | National | 0.74 |
| 23th Mar | 18 | Reduce importations | 14 | National | 0.92 |
| 5th Mar | 0 | Social distancing | 7 | National | 0.64 |
| | 0 | Stay at home work | 7 | National | 0.64 |
| | 0 | Stay at home other | 7 | National | 0.64 |
| 12th Mar | 7 | Social distancing | 7 | National | 0.90 |
| | 7 | Stay at home work | 7 | National | 0.66 |
| | 7 | Stay at home other | 7 | National | 0.90 |
| 19th Mar | 14 | Stay at home work | 14 | National | 0.72 |

Table 5: Detail of interventions implemented in simulations for Spain. "Day" measures the number of days from the 5th of March 2020. "Delay" is the time interval during which the intervention increases linearly to the "Target" value: a value of zero means the intervention applies immediately. "Extent" refers to the application of the intervention on the Spanish territory: all interventions for Spain are"National" i.e. they apply to the whole Spanish territory. Values for social distancing and stay-at-home policies are derived by Google data.

of the rates as well as a discussion of their reliability and a projection of worldwide deaths for an unrestricted epidemic.

#### A.6 Fitting procedure

We use two approximate Bayesian computation fitting procedures to estimate model parameters. The first is based on Sequential Monte Carlo, while the second is based on a description via Gaussian Processes. The second method has the advantage of being particularly efficient, and is thus useful to our modelling approach.

##### A.6.1 Sequential Monte Carlo

In approximate Bayesian computation based on Sequential Monte Carlo [24], a number of sampled parameter values (called particles) are propagated through a sequence of intermediate distributions that increasingly approach a target distribution. This is achieved by associating to each intermediate distribution an error $\epsilon_j$ which is reduced for increasing $j$. The algorithm generates particles at each iteration: each particle is a set of parameters for which $B_t$ simulations of disease transmission are executed. At the end of each simulation the resulting set of disease incidence estimates is compared with the original dataset of incidence of the disease, while $0 \leq b_t \leq B_t$ counts the number of simulations that ended at distance less than $\epsilon_j$. Each particle gets an associated weight measuring its relevance. Each generation, the set of accepted particles is sampled according to weights $w_j$, and perturbed by a perturbation kernel $\mathcal{K}$ in order to produce a new set of $\mathcal{N}$ particles.

The weights are defined as [24]:

$$w_j = \begin{cases} 1, \\ \dfrac{b_t \pi(q_t)}{\sum_{j=1}^{\mathcal{N}} w_{t-1} \mathcal{K}_j(q_{t-1}, q_t)} \end{cases}$$

The case $B_t = 1$ is used when transmission is obtained from deterministic models, while for stochastic models a value $B_t > 1$ is expected. Consequently, the fitting of stochastic models is lengthy as several runs are required for each set of parameters in order to estabilish the relative goodness of different sets of parameters.

The fitting algorithm however requires knowledge of the distance of the time series produced by the simulation to the original data. Since this information is richer than a simple acceptance status, we include this information in the weights and limit the number of simulations for each parameter to $B_t = 1$. Our weights are thus defined as:

$$w_j^t = \begin{cases} \dfrac{\epsilon_t}{\delta_j^t}, \\ \dfrac{\epsilon_t \pi(q_t)}{\delta_t \sum_{j=1}^{\mathcal{N}} w_{t-1} \mathcal{K}_j(q_{t-1}, q_t)} \end{cases}$$

When particles produce time series very close to the data, then the ratio $\delta_j^t / \epsilon_t$ is small and the weights get amplified. We use an adaptive sequence of tolerances where $\epsilon^{t+1}$ is the $\eta_0$ quantile of the distances $d_0^t, \ldots, d_N^t$ of accepted particles at generation time $t$, and $\eta_0$ is a tunable parameter that we set to $\eta_0 = 0.4$.

The distance from a simulated time series $\{e_j\}$ and the corresponding data set $\{d_j\}$ is evaluated using the following function:

$$\delta = \begin{cases} 0 & \text{for} \quad e_j = d_j = 0 \\ \displaystyle\sum_j \frac{|e_j - d_j|^2}{e_j + d_j} & \text{otherwise} \end{cases} \tag{5}$$

The above expression can be used when both data and estimates are close to zero in the early stage of the outbreak: the expression avoids divergence when either $e_j$ or $d_j$ are zero. Also, (5) is continuous when both $e_j$ and $d_j$ are zero.

##### A.6.2 Parallel implementation

The fitting procedure runs several simulations (each using parallel computation) in parallel, thus creating a multi-level parallel simulation. The simulations run independently so that if one takes longer it will not delay the others, and they only synchronize when enough simulation succeed to produce a new generation of particles. In order to achieve this, one of the parallel processes uses an extra thread to control the independent parallel simulations during the fitting procedure and to keep track of how many were successful. When the target number of successful simulations is achieved, it synchronizes them so that the main process can collect all information, produce a full new set of particles and distribute it back. Each simulation has a dedicated communication channel where information is exchanged, whilst communication between simulations and the control thread is handled by a separate channel. Both the thread and the main process may communicate with other processes depending on the kind of messages, and a synchronization mechanism between the thread and the main process, which activates when a new generation of particles is produced, ensures that the two do not end up in deadlock and that the thread can be safely terminated upon completion.

##### A.6.3 Bayesian Optimization for Likelihood Free Inference

Bayesian Optimization for Likelihood Free Inference is an approach (detailed in Gutmann and Corander [25] and in Järvenpää [26]) for estimating the posterior distribution of a model parameters usable with expensive simulations as it reduces the number of executions required for parameter estimation. The method is based on a description of the relation between model parameters and the summary statistics, in our case defined by (5), with a Gaussian Process. Given a set of parameter values $\{\boldsymbol{\theta}_1, \ldots, \boldsymbol{\theta}_t\}$ and a set of distances $\{f_1, \ldots, f_t\}$, the posterior distribution is a Gaussian distribution with mean $\mu(\boldsymbol{\theta})$ and variance $v(\boldsymbol{\theta}) + \sigma_n^2$, with

$$\mu(\boldsymbol{\theta}) = m(\boldsymbol{\theta}) + \mathbf{k}_t(\boldsymbol{\theta})^\intercal \mathbf{K}_t^{-1}(\mathbf{f}_t - \mathbf{m}_t).$$

The covariance matrix $\mathbf{K}_t$ has the form:

$$K_{t,ij} = k(\boldsymbol{\theta}^{(i)}, \boldsymbol{\theta}^{(j)}) + \delta_{ij}\sigma_n^2$$

where $\delta_{ij}$ is the Kronecker delta, and $k(\boldsymbol{\theta}, \boldsymbol{\theta}')$ describes a squared exponential covariance function

$$k(\boldsymbol{\theta}, \boldsymbol{\theta}') = \sigma_f^2 \exp\left(-\frac{1}{2}\sum_q \frac{(\theta_q - \theta'_q)^2}{\lambda_q^2}\right)$$

and

$$\mathbf{k}_t(\boldsymbol{\theta}) = \{k(\boldsymbol{\theta}, \boldsymbol{\theta}^{(1)}), \ldots, k(\boldsymbol{\theta}, \boldsymbol{\theta}^{(t)})\}^\intercal$$

We observe that in our work, the distance is a non-symmetric function of parameters: figure 3 shows the dependency of the distance from the parameter in a simplified case were we fit one parameter on a set of data produced by our simulations. The stochastic fluctuations around the mean are small: in order to reproduce the correct dependency of $m(\boldsymbol{\theta})$ on the parameters, we assume a polynomial form of order higher than two. As shown in figure 3, a quartic function fits the data extremely well. In multiple dimensions, cross term parameters are expected due to the influence of some parameters (in particular those describing the initial condition) on the others. A full quartic function in multiple dimensions would require a large number of parameters: we simplify its form as:

$$m(\boldsymbol{\theta}) = a + \sum_p b_p\theta_p + \sum_p\sum_{q\geq p} c_{pq}\theta_p\theta_q + \sum_p\sum_q d_{pq}\theta_p^2\theta_q + \sum_p\sum_{q\geq p} e_{pq}\theta_p^2\theta_q^2$$

where we are essentially neglecting ternary terms. The optimal set of hyperparameters $\{a, b_q, c_{pq}, d_{pq}, e_{pq}\}$ describing the mean function $m(\boldsymbol{\theta})$ that minimizes $S_t = |\mathbf{m}_t - \mathbf{f}_t|^2$ can be found using a minimization or a least squares algorithm, and the remaining hyperparameters are found by maximizing the log marginal likelihood:

$$\log P(\mathbf{y}|\boldsymbol{\theta}) = -\frac{1}{2}\mathbf{y}^\intercal \mathbf{K}_t^{-1}\mathbf{y} - \frac{1}{2}\log|\mathbf{K}_t| - \frac{n}{2}\log 2\pi$$

with $\mathbf{y} = \mathbf{f}_t - \mathbf{m}_t$.

The threshold for acceptance is selected as the $\epsilon_t$-quantile of the distribution of distances $\{\delta_j\}_t$. The acquisition rule used to extend the set of observations in the Gaussian Process

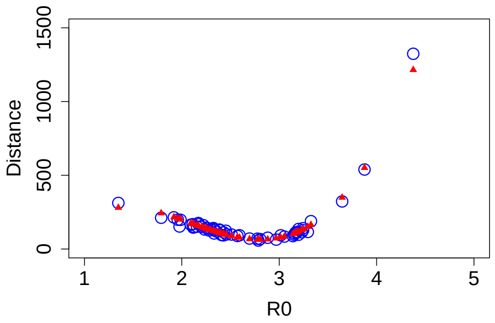


Figure 3: The relation between distance and parameters in the simplified case of one parameter. Data were obtained by fitting a synthetic output from a simulation produced with $R_0 = 2.7$. The simulation uses a map of Italy with a population homogeneously reduced to one tenth for speed. The blue circles are raw distances between simulation outputs generated during the fitting process and the synthetic data set. The red triangles correspond to the values of the mean function $m(\boldsymbol{\theta})$ fitted to the data, assuming a quartic form for the dependence.

selects the next point by extracting it from the distribution $\pi^2(\boldsymbol{\theta})\mathbb{V}(\boldsymbol{\theta})$ (method "rand_maxvar" in Ref. [26]) using a Metropolis-Hastings algorithm, where $\mathbb{V}(\boldsymbol{\theta})$ is the variance of the posterior ($\epsilon$ dependent) probability distribution that has the form of eq. 10 in Ref. [26], taking into account that $m_{1:t}(\boldsymbol{\theta})$ should be replaced by $\mu(\boldsymbol{\theta})$. The posterior distributions for each parameter as well as the corresponding time series are obtained by sampling the expected probability distribution in eq. 9 of Ref. [26] (substituting $m_{1:t}(\boldsymbol{\theta})$ with $\mu(\boldsymbol{\theta})$) using a Metropolis-Hastings algorithm.

##### A.6.4 Initial condition during fitting

All simulations show that an initial detected infection produces a large number of undetected cases, whereas a new detected infection only occurs after a considerable delay. It would be unable thus to explain the rapid increase in cases observed in Italy and in Kenya. In order to approximate the initial growth of the number of detected cases, we assume that a continuous import of cases is active before day zero. During this "burn-in" phase, a non-zero importation rate grants new imported cases in the country. To take into account the possibility that undetected cases may have entered the country, we assume that imported cases enter the exposed class and may thus become either detected or undetected cases. The importation rate is tuned according to the local data available.

To implement a spatially dependent importation rate, we use the following method. Let $\omega_{jk}$ be the importation rate in age group $j$ and county $k$, we may write it down as:

$$\omega_{jk} = f_{jk}\ \omega^0 \tag{6}$$

were $f_{jk}$ is the estimated ratio of cases in age group $j$ and location $k$. This may be obtained, for instance, by dividing the number of cases in a given area by the corresponding population size. If the number of cases is not age stratified, then cases occur in all age groups proportionally to the group age size. Potential importation thus occur at rate $\omega^0$, however each importation events is accepted only with a probability $f_{jk}$. To maximize the efficiency, we define $\omega^i = \max_{jk} f_{jk}$ so that the expression may be rewritten as

$$\omega_{jk} = \tilde{f}_{jk}\ \omega^i \omega^0$$

Now, potential importation may occur at rate $\tilde{\omega} = \omega^i \omega^0$, which may be estimated through fitting, however the acceptance probability is maximized to $\tilde{f}_{jk} = f_{jk}/\omega^i$, and takes value 1 in at least one age group and county.

We assume that importations start flowing in the country a number of days $\tau$ before day zero, with $\tau$ a fittable parameter. This "burn-in" phase does not stop at day zero, but continues throughout the simulation and may be modulated and eventually reduced to zero by using the appropriate interventions. We tuned the importation rate according to governamental interventions and to flight data reported in Ref. [27]. Further data on passenger reduction during the COVID pandemic is available on a number of newspapers and news outlets, for instance [28, 29, 30, 31, 32, 33].

##### A.6.5 Estimated parameters and priors

The simulations uses information available from the literature on various parameters as detailed in Table 1, whilst a few more are estimated by the fitting procedure. Table 6 shows the parameters estimated, their priors and the corresponding range of values.

#### A.7 Susceptibilities

Estimation of susceptibilities via the simulation model is exceptionally time consuming and hard, given that we need to fit a total of 12 parameters (if we fix the susceptibility of one of the age groups, the 80+, to 1 and fit concurrently $R_0$, $\gamma$, $t_0$ and $\omega$). We thus follow an approximated approach. Let $K_{ij}$ be the age-mixing matrix, the force of infection is given by

$$\frac{R_0\gamma}{\lambda_{\tilde{K}_{\mathrm{CH}}}} \sum_j \tilde{K}_{ij} \frac{I_j + U_j}{N_j} \tag{7}$$

| Parameter | Symbol | Prior |
|---|---|---|
| Basic reproductive number | $R_0$ | Uniform$[1\ ,\ 8]$ |
| Recovery rate (inverse) | $1/\gamma$ | Uniform$[0.2,\ 8]$ |
| Duration of "burn-in" phase | $\tau$ | Uniform$[0,\ 60]$ |
| Importation rate (logarithm) | $\log\omega$ | Uniform$[\log(10^{-5}),\ \log(1)] + \log\omega^p$ |

Table 6: Fitted parameters and priors. All priors are uniformly distributed: the factor $\omega^p$ appearing in the prior for $\omega$ is a parameter which shifts the range of the uniform distribution to adapt it to a specific country. This modulating term may be tuned to adapt the prior to the specific temporal duration and the size of the area that each $f_{jk}$ term in 6 refers to.

where $\tilde{K}_{ij} = \sigma_i K_{ij}$ is an age-mixing transmission matrix that takes into account the age dependent susceptibilities. The term $\lambda_{\tilde{K}_{\mathrm{CH}}}$ is the maximum eigenvalue of the Chinese age-mixing transmission matrix calculated using the same set of susceptibilities $\{\sigma_i\}$. In this formulation, the susceptibilities do not affect $R_0$, and we can thus use them to modulate the age-distribution of the expected number of cases.

The age distribution of the infectives is given by the components of the eigenvector corresponding to the maximum eigenvalue of the matrix $\tilde{K}_{ij}N_i/N_j$. However, the contact matrix $K_{ij}$, and thus the transmission matrix $\tilde{K}_{ij}$, is modified during the dynamics in a nontrivial way: we can express the fact that it changes during the simulation by explicitly stating its time dependence $\tilde{\boldsymbol{K}}(t)$. We may however find an approximated matrix $\tilde{\boldsymbol{C}} \simeq \frac{1}{T}\int_T \tilde{\boldsymbol{K}}(t)\ dt$ that would act as an effective transmission matrix during the time interval $T$. Since the major modification of the contact patterns comes from the selective alterations of the contribution of the four components, we can estimate $\tilde{\boldsymbol{C}}$ for a reference simulation with $\sigma_i = 1 \quad \forall i$, by finding the linear combination of the four components (home, work, school, other) that, combined with ad-hoc hyper-susceptibilities, reproduces its age distribution of cases:

$$\tilde{\boldsymbol{C}} = \sigma_i^0 \left( K_{ij}^{\mathrm{home}} + \alpha_1 K_{ij}^{\mathrm{work}} + \alpha_2 K_{ij}^{\mathrm{school}} + \alpha_3 K_{ij}^{\mathrm{other}} \right)$$

where $\sigma_9^0 = 1$ and the remaining hyper-parameters $\{\alpha_1, \alpha_2, \alpha_3, \sigma_1^0, \cdots, \sigma_8^0\}$ can be fitted using a standard ABC-SMC algorithm. The optimal value of hyper-parameters defines a transmission matrix $\tilde{\boldsymbol{C}}$ where the eigenvector corresponding to its largest eigenvalue, describes the age distribution of cases produced by the reference simulation.

We then use the resulting matrix $\tilde{\boldsymbol{C}}$ to find the best set of $\sigma_i$ that solves the eigenvalue equation

$$\sigma_i \tilde{C}_{ij} v_j = \lambda v_i$$

producing a set of $v_i$ such that $a_i v_i$ is close to the distribution of cases observed in data, with $a_i$ being the ascertainment rate in the main text.

#### A.8 Data sources

The model accepts as input daily cases or cumulative daily cases, daily deaths or cumulative daily deaths: the fitting procedure can be instructed to fit cumulative or non cumulative daily cases and/or deaths. This work fits simulations to daily deaths. In addition to the COVID-19 Data Repository by the Center for Systems Science and Engineering at Johns Hopkins University (JHCSSE) [34, 35], data is available through national surveillance data bases. Italian data is available from the Italian Protezione Civile [36], Spanish data is available through daily reports from the Gobierno de España [37]. UK data is available through a dedicated UK government website [38]. It is worth noting that a few countries (Spain and the UK among those discussed in this work) have modified the definition of case and/or death at some point during the pandemic: for the UK, data has been modified retroactively, whilst for Spain it has not. Despite such changes would hardly modify the overall framework and results, we limited our fitting procedure to time span that excludes major changes, and committed to JHCSSE data set.

For the initial condition, we used the goegraphical detailed reported number of cases. This kind of local data is available in different formats for different countries. For Italy, the daily number of cases per province is available from the Protezione civile [36]: there are 110 provinces in Italy, providing a spatially detailed description, but no information on the age distribution of cases is available. Spain reported the daily number of cases per Autonomous Community: there are 16 Autonomous Communities in Spain if we exclude the Autonomous Communities located in Africa (Ceuta, Melilla and the Canary Islands): no information on the age distribution of cases is available. For the UK, the situation is more complex as the four constituting nations have separate databases for local cases. England and Wales provide data on daily incidence by Local Authority [38]: there are 380 Local Authorities, providing a detailed spatial description. Age distribution of cases is also available, but at the level of NHS board, which are extended areas covering large parts of the two nations: there are 9 NHS areas. For Wales, the daily number of cases per Local Government is available from the Welsh Government [39]. Northern Ireland is handled as a single entity and not partitioned in its component constituencies, thus we use data from the main UK website [38]. For Scotland, we used data per Scottish NHS board [40]. This data however was incomplete as the exact number of cases in each board was unknown if it was less than five cases, which occurred during the initial phases of the Scottish outbreaks. We thus ended up assigning cases based on population size. Note however that, currently, historical daily data is available at the level of Local Authorities on the Scottish Government website [41]. In all countries we selected a date representative of the initial distribution of cases and used the spatial case distribution to tune the importation rate: this was typically close to the date that the first deaths were reported.

#### A.9 Implementation structure

The main simulation code, stored in the `Master` directory, uses a simulation engine wrote by one of the authors to simulate the spatio-temporal dynamics of the disease. The engine takes a file specifying the compartmental model structure, general simulation parameters and other general details and produces an executable that can be submitted to a computer cluster to perform the simulation. Due to the complexity of the model, this file is generated by a python script called `generate.py`. This script imports country-specific parameters described in the `config.py` file. Code implementing features beyond the compartmental model are detailed in a C++ file called `lse-userdefined.cpp`. This file is also in part generated by the `generate.py` script by adding code to a base `lse-userdefined-base.cpp` file. This code is responsible for the handling of all model specific features like contact tracing, interventions, etc. This file imports country specific code from coutry specific files *Country*`-setup.cpp`, that detail what intervention a country has implemented and how they were implemented.

In addition to the main simulation code, additional programs perform the data reshaping of the WorldPop maps. Code stored in the `Master/Setup` directory is responsible for producing coarse-grained maps with a given resolution in km from the original WorldPop age stratified maps. Some utilities of the simulation engine are used in the `Map` directories to build gridded maps of county identifiers based on shapefiles describing the geometry of administrative divisions for the country, and to produce partition maps that are used by the engine to distribute the workload evenly among computing nodes. Code within the `Map/Mobility` directories is used to estimate parameters for the human mobility model based on the data available on movements between areas of the country: these may or may not coincide with the administrative subdivisions of the country and are stored in specific shapefiles.

An installation file `install.sh` in the root directory is responsible to automatically prepare for simulating a specified country: it automatically downloads data from the WorldPop database, runs the code that builds the age stratified coarse grained maps, produces partition files and sets up the simulation code for estimating human mobility parameters and running the main simulation code. Human mobility parameters are not estimated automatically after installation as this may be lengthy: estimated values are already pre-stored in the configuration files. The code is available on github at `https://github.com/andreaparisi-science/SpatialCOVID19`

[18] https://www.ine.es/dyngs/INEbase/es/operacion.htm?c=Estadistica_C&cid=1254736176952&menu=ultiDatos&idp=1254735572981

[24] Toni, T., Welch, D., Strelkowa, N., Ipsen, A., and Stumpf, M.P.H. (2009). Approximate Bayesian computation scheme for parameter inference and model selection in dynamical systems. Journal of The Royal Society Interface 6, 187202.

[34] Dong, E., Du, H., and Gardner, L. (2020). An interactive web-based dashboard to track COVID-19 in real time. The Lancet Infectious Diseases 20, 533534.

[35] COVID-19 Data Repository by the Center for Systems Science and Engineering (CSSE) at Johns Hopkins University. https://github.com/CSSEGISandData/COVID-19.

[36] Consiglio dei Ministri, Dipartimento della Protezione Civile: Emergenza Coronavirus. Dati COVID-19 Italia: Monitoraggio situazione. https://github.com/pcm-dpc/COVID-19/blob/master/dati-andamento-nazionale/dpc-covid19-ita-andamento-nazionale.csv. Retrieved on the 15th of May 02020

[37] Gobierno de España, Ministero de Sanidad, Centro de Coordinación de Alertas y Emergencias Sanitarias. Enfermedad por el coronavirus (COVID-19). Actualización no 31-Actualización 108. url-https://www.mscbs.gob.es/profesionales/saludPublica/ccayes/alertasActual/nCov/situacionActual.htm

[38] UK Government, Public Health England and NHSX. Coronavirus (COVID-19) in the UK. https://coronavirus.data.gov.uk/cases. Retrieved in August 2020.

[39] Public Health Wales. Coronavirus (COVID-19). https://phw.nhs.wales/topics/latest-information-on-novel-coronavirus-covid-19/. Coronavirus data dashboard: https://public.tableau.com/profile/public.health.wales.health.protection#!/vizhome/RapidCOVID-19virology-Public/Headlinesummary, retrieved August 2020.

[40] Scottish Government. Coronavirus (COVID-19): trends in daily data. https://www.gov.scot/publications/coronavirus-covid-19-trends-in-daily-data/ Retrieved August 2020.

[41] Daily COVID-19 Cases in Scotland. Daily Case Trends By Council Area. https://www.opendata.nhs.scot/dataset/covid-19-in-scotland/resource/427f9a25-db22-4014-a3bc-893b68243055
