## Supplementary Material SM2 for "Spatially resolved simulations of the spread of COVID-19 in European countries"

Determination of detection rates, hospitalizations, ICU admissions, death rates of severe and critical cases from Spanish data sets. Comparison with other countries.

The Spanish Government has published detailed age stratified data on hospital and critical care admissions. I looked into the data to get sensible estimates of hospitalization. It happens that the Spanish serological survey also came out, age stratified and quite detailed. Using these, and additional information from a published article of ICU admissions in Italy, one can derived all relevant rates/probabilities and, through comparison with other countries, say something on the reliability of the data.

### Spanish population pyramid

Get age pyramid for Spain (5 years age groups) these estimates are produced via processed WorldPop data (2020 estimates)

```
age.pyram.17 <- as.matrix( read.table("Data/Spain_5km_stats.dat",
                                      header=FALSE) )
age.pyram.17 <- unlist(age.pyram.17[,3])
```

Build reduced age pyramid (10 years age groups) by aggregating

```
get.10y.from.5y <- function( ap.17 )  {
    ap.9 = c()
    for (jj in seq(1,16,2))  ap.9 <- cbind(ap.9, ap.17[jj]+ap.17[jj+1])
    ap.9 <- cbind(ap.9, ap.17[17])
}

get.5y.from.10y <- function( ap.9, bp.17 )  {
    ap.17 = c()
    for (ii in 1:8)  {
        ratio <- bp.17[(2*ii-1):(2*ii)]
        ratio <- ratio[1]/(ratio[1]+ratio[2])
        ap.17 <- c(ap.17, ap.9[ii]*ratio, ap.9[ii] - ap.9[ii]*ratio)
    }
    ap.17 = c(ap.17, ap.9[9] )
}

age.pyram.9 <- get.10y.from.5y( age.pyram.17 )
```

### Spanish survey data

Take data from Spahish survey: number of tested and percentage of positive

```
# Data from spanish survey
spanish.survey.count.20 <- c(268, 1693, 2857, 3425, 3221, 2805, 2606, 3050, 4000,
                             5174, 5330, 5263, 5187, 4560, 3568, 2931, 2161, 1410,
                             968, 420)
```

```
spanish.survey.perc.20  <- c(1.1, 2.2, 3.0, 3.9, 3.8, 4.5, 4.8, 3.8, 4.6,
                             5.3, 5.7, 5.8, 6.1, 5.9, 6.2, 6.9, 6.1, 5.1, 5.6, 5.8)
```

Estimate the number of positives in survey

```
spanish.survey.posit.20 <- spanish.survey.count.20 * spanish.survey.perc.20 / 100.0
```

Reduce survey data to 17 age groups

```
reduce.from.20.age.groups <- function( data )  {
    resp <- data[2:18]
    resp[1] <-resp[1] + data[1]
    resp[17] <- resp[17] + data[19] + data[20]
    return(resp)
}

spanish.survey.count.17 <- reduce.from.20.age.groups( spanish.survey.count.20 )
spanish.survey.posit.17 <- reduce.from.20.age.groups( spanish.survey.posit.20 )
```

Reduce data of spanish survey to 10y age groups

```
spanish.survey.count.9 <- get.10y.from.5y( spanish.survey.count.17 )
spanish.survey.posit.9 <- get.10y.from.5y( spanish.survey.posit.17 )
```

Estimate positive counts on national basis using the 5 years age groups

```
spanish.survey.perc.9 <- spanish.survey.posit.9 / spanish.survey.count.9
spanish.estimate.count.9 <- spanish.survey.perc.9 * age.pyram.9
print(paste('Check: 5% ~', sum(spanish.estimate.count.9)/(sum(age.pyram.9))*100.0) )
```

```
## [1] "Check: 5% ~ 5.02894653153549"
```

#### Admission to ICU and overall (severe + critical) death rates

Information from Spain at 16.05.2020 - 10 years age groups. This is age stratified information on 239,125 cases, and includes hospital admissions, ICU admissions (from hospitalized), and deaths (both ICU and hospital, for a total of 19,186 deaths). The age stratified information is a subset of the total cases (about 70%) for which information on sex and age is available.

```
cases.total.9 <- c(877, 1637, 13461, 22639, 35135, 42794, 34360, 32443, 37463+18316)
cases.hosp.9 <- c(279, 277, 1477, 3811, 8666, 14075, 17264, 20794, 18842+6346)
cases.ICUs.9 <- c(39, 23, 89, 274, 740, 1562, 2534, 2261, 332+56)
cases.deaths.9 <- c(2, 5, 23, 63, 201, 611, 1695, 4632, 7872+4082)
```

The same report provides further information on all cases, setting the total death toll to 27,650. Using this information, we rescale the deaths to account for the true death toll. This is supported by the fact that the rate of hospitalization and ICU admission for the wider set is similar to that for the age stratified set.

```
total.deaths <- 27650
cases.deaths.9 <- cases.deaths.9*total.deaths/(sum(cases.deaths.9))
```

Determine ascertainment rate. This is the rate of detection of cases, not asymptomatic rates. This is given by the count of detected cases divided by the count of totally infected plus the dead individuals (which are there but would not appear in the count of individuals affected by COVID19). We get the rates for 10y age groups.

```
ascert.rate.9 <- cases.total.9 / (spanish.estimate.count.9+cases.deaths.9)
print(ascert.rate.9)
```

```
##              [,1]        [,2]       [,3]       [,4]       [,5]       [,6]
## [1,] 0.006781274 0.007714451 0.05492951 0.08193389 0.07342566 0.09102403
##             [,7]      [,8]      [,9]
## [1,] 0.09354722 0.1070177 0.2893215
```

### ICU death rates using Italian data

Estimates of ICU deaths are based on a study published recently on about 1600 patients admitted to ICU in Lombardy, Italy [Grasselli, G., et al. (2020). Baseline Characteristics and Outcomes of 1591 Patients Infected With SARS-CoV-2 Admitted to ICUs of the Lombardy Region, Italy. JAMA 323, 1574–1581]. The patients in the study were admitted between the 20th of February and the 18th of March (28 days), and the study reports on their status on the 25th of March (35 days).

Copy of Italian data: (n) -> number of cases, (d) -> number of deaths, (s) -> number of active cases

```
# Derive estimated critical death rate from Italian data
ita.ICUn.9 <- c(0, 2,  0, 56, 142, 423, 596, 340, 22)
ita.ICUd.9 <- c(0, 0,  0,  4,  16,  63, 174, 136, 12)
ita.ICUs.9 <- c(0, 2,  0, 32,  91, 270, 353, 164,  8)
```

Estimate must take into account that a fraction of those still in hospital will die. Assume the usual $dI/dt$ for an equivalent SIR model, with $R_0(t)$ linearly decreasing between $R_0 = 3$ and 1 [Riccardo, F., et al. (2020). Epidemiological characteristics of COVID-19 cases in Italy and estimates of the reproductive numbers one month into the epidemic. MedRxiv 2020.04.08.20056861.]. This implies $I(t) \propto \exp(\int_0^t (R_0(\tau) - 1)\,\gamma\,d\tau)$.

$$
\begin{gathered}
R_0(t) = R_0 + (1 - R_0)t/T \quad \text{where } R_0 = 3, \text{ and } T = 28 \text{ days} \\
\frac{dI}{I} = (R_0(t) - 1)\gamma\,dt = (R_0 - 1)\,(1 - t/T)\,\gamma\,dt \\
\Rightarrow \\
I(t) \propto \exp\left((R_0 - 1)t\,(1 - t/2T)\,\gamma\right)
\end{gathered}
$$

First build a function that returns $I(t)/I(0)$, then a function that returns the entry time of n severe admissions to ICU. Use simple rejection method to draw from the normalized I(t). Use $\gamma = 1/7$: this is the approximated equivalent SIR model.

```
epigrowth <- function(t, R0, gamma)  {
#   alpha = (R0-1)*gamma
#   return(exp(alpha*t)/alpha)
    return( exp( (R0-1)*t*(1-t/(2*28))*gamma ) )
}

# Returns numbers distributed according to I(t)/I(0). These are hospitalization times
repigrowth <- function( n, R0, gamma )  {
    maxobj <- optimize(function(x) {return(epigrowth(x, R0, gamma))},
                       interval=c(0, 28), maximum=TRUE)
    x.max <- maxobj$maximum
    y.max <- maxobj$objective
    y.min <- 1
    theta = y.max - y.min
    vec <- c()
    for (jj in 1:n)  {
        repeat {
            x = runif(1)*28
            y = runif(1) < epigrowth(x, R0, gamma)/theta
            if (y)  {
```

```
                break
            }
        }
        vec <- c(vec, x)
    }
    return(vec)
}
```

Search for death rate that produces the expected deaths by the 25th of May. The final number of deaths in ICU will be higher than the number of deaths registered up to the 25th.

```
estimate.ICU.deaths <- function(rate, age)  {
    vals <- c()
    nn = floor(ita.ICUn.9[age]*rate)
    if (nn > 0)  {
        for (jj in 1:1000)  {
            t.to.death.ITA <- rpois(nn, 7)
            t.hospital.ITA <- repigrowth(nn, R0=3.0, gamma=1.0/7.0)
            # From 20-02-2020 to 18-03-2020 (included)
            vals <- cbind( vals, sum(t.to.death.ITA+t.hospital.ITA <= 35) )
        }
    } else {
        vals = 0
    }
    return( vals )
}


findRate <- function(rate, age)  {
    return( mean(estimate.ICU.deaths(rate, age)) - ita.ICUd.9[age] )
}


pg1 <- txtProgressBar(min=1, max=length(ita.ICUn.9), style=3)
critic_to_death.9 <- c()
for (age in 1:length(ita.ICUn.9))  {
    setTxtProgressBar(pg1, age)
    rate <- uniroot(findRate, c(0,1), age)$root
    critic_to_death.9 <- cbind(critic_to_death.9, rate)
}
```

```
##
  |
  |                                                                  |   0%
  |
  |========                                                          |  12%
  |
  |================                                                  |  25%
  |
  |========================                                          |  38%
  |
  |================================                                  |  50%
  |
  |=========================================                         |  62%
  |
```

```
  |==================================================                |  75%
  |
  |==========================================================        |  88%
  |
  |==================================================================| 100%
```

```r
print( critic_to_death.9 )
```

```
##      rate rate rate       rate      rate      rate      rate      rate
## [1,]    0    0    0 0.08922351 0.1267597 0.1631505 0.3187646 0.4354254
##           rate
## [1,] 0.6363434
```

```r
print( paste('Check:',sum(critic_to_death.9*ita.ICUn.9),'>',sum(ita.ICUd.9) ) )
```

```
## [1] "Check: 444.036950114709 > 405"
```

```r
print(critic_to_death.9*ita.ICUn.9)
```

```
##      rate rate rate     rate     rate     rate     rate     rate     rate
## [1,]    0    0    0 4.996517 17.99987 69.01266 189.9837 148.0446 13.99955
```

### Finalizing data

Estimate deaths in ICUs and hospital in Spain using Italian rates. Estimate death rates for severe cases

```r
ICUs.deaths.9 <- (cases.ICUs.9 * critic_to_death.9)
hosp.deaths.9 <- cases.deaths.9 - ICUs.deaths.9
# Death rates from non-ICU cases
hosp_to_death.9 <- hosp.deaths.9 / (cases.hosp.9-cases.ICUs.9)
hosp_to_ICUs.9 <- cases.ICUs.9 / cases.hosp.9
```

Hospitalization rates are the crude rates, given cases.total is the number of detected cases.

```r
ascert_to_hosp.9 <- (cases.hosp.9-cases.ICUs.9)/cases.total.9
ascert_to_ICUs.9 <- cases.ICUs.9/cases.total.9
print("Sympt to hosp")
```

```
## [1] "Sympt to hosp"
```

```r
print(ascert_to_hosp.9)
```

```
## [1] 0.2736602 0.1551619 0.1031127 0.1562348 0.2255870 0.2924008 0.4286962
## [8] 0.5712480 0.4446118
```

```r
print(ascert_to_ICUs.9)
```

```
## [1] 0.044469783 0.014050092 0.006611693 0.012103008 0.021061619 0.036500444
## [7] 0.073748545 0.069691459 0.006956023
```

```r
print(cases.hosp.9/cases.total.9*hosp_to_ICUs.9)
```

```
## [1] 0.044469783 0.014050092 0.006611693 0.012103008 0.021061619 0.036500444
## [7] 0.073748545 0.069691459 0.006956023
```

```r
print("The last two should be the same")
```

```
## [1] "The last two should be the same"
```

Output all rates, assuming 5y age groups (simulation ready... more or less: see at the end)

```r
ascert_to_death.9 <- ascert_to_hosp.9*hosp_to_death.9 + ascert_to_ICUs.9*critic_to_death.9
dat <- data.frame( age=seq(0,8*10, by=10), arate=c(ascert.rate.9),
                   a2s=ascert_to_hosp.9, a2c=ascert_to_ICUs.9,
                   s2d=c(hosp_to_death.9), c2d=c(critic_to_death.9) )
#write.csv(dat, file="../../Data/Other/rates.csv", row.names=FALSE)
tail( dat, n=10 )
```

```
##   age       arate       a2s         a2c        s2d        c2d
## 1   0 0.006781274 0.2736602 0.044469783 0.01200963 0.00000000
## 2  10 0.007714451 0.1551619 0.014050092 0.02836919 0.00000000
## 3  20 0.054929514 0.1031127 0.006611693 0.02388081 0.00000000
## 4  30 0.081933888 0.1562348 0.012103008 0.01875757 0.08922351
## 5  40 0.073425664 0.2255870 0.021061619 0.02471234 0.12675967
## 6  50 0.091024028 0.2924008 0.036500444 0.05000437 0.16315049
## 7  60 0.093547221 0.4286962 0.073748545 0.11099852 0.31876460
## 8  70 0.107017749 0.5712480 0.069691459 0.30707026 0.43542543
## 9  80 0.289321537 0.4446118 0.006956023 0.68470426 0.63634337
```

The interesting aspect of these rates is that they are not strictly dependent on the knowledge of the detection rate. In other words, sympt.rate.9*symp__to__hosp.9 is the rate an average infected individual will be hospitalized (severe case), independently on being symptomatic or asymptomatic. That means that as long as we have a detection rate, or better a symptomatic rate that is compatible with (meaning greather than) the hospitalization rate, we can rescale one of the terms of the multiplication according to what the other term becomes. What is invariant is the product. By splitting the product into detection rate and hospitalization rate, we retain the information on the detection rate: this could be used for comparison between countries.

### Comparisons and checks

We can compare the resulting rates with with the Infectious Fatality Ratio (IFR) from the Verity paper [Verity, R., et al. (2020). Estimates of the severity of coronavirus disease 2019: a model-based analysis. The Lancet Infectious Diseases.], and with other countries. This will provide an insight into the reliability of the estimates obtained above.

#### Comparing with Verity IFR

How do expected deaths for Spain based on above numbers compare with those predicted with Verity IFR?

```r
expected.deaths.9 <- age.pyram.9*ascert.rate.9*(ascert_to_hosp.9*hosp_to_death.9+
                                                ascert_to_ICUs.9*critic_to_death.9)
print(sum(expected.deaths.9))
```

```
## [1] 454626.9
```

```r
IFR <- c(0.00161, 0.00695, 0.0309, 0.0844, 0.161, 0.595, 1.93, 4.28, 7.80)/100.0
print(sum(age.pyram.9*IFR))
```

```
## [1] 633294.1
```

Here we are using a rough estimate of the number of deaths if 100% of the population were affected. The Verity IFR predicts a higher death toll, about 1.5 times higher It is worth exploring where the difference comes from

```r
#expected.deaths.9 <- get.10y.from.5y( expected.deaths.17 )
print((age.pyram.9*IFR-expected.deaths.9)/expected.deaths.9)
```

```
##             rate    rate     rate     rate     rate     rate     rate
## [1,] -0.2776071 1.04667 1.284502 1.568531 1.659569 2.176815 1.902012
##           rate       rate
## [1,] 0.9437022 -0.1271078
```

```r
print(sum(age.pyram.9*IFR))
```

```
## [1] 633294.1
```

```r
print(sum(expected.deaths.9))
```

```
## [1] 454626.9
```

The general consensus is that death in Spain (as in many European countries) are underestimated as they typically miss many elderly dying home or in care homes with typical COVID19 symptoms. However, here the bulk of the difference does not come from the oldest age groups, but from middle aged groups. This poses a doubt, which requires comparison with other countries.

We can have a look at two other data sets from European countries: from Italy and Germany.. At the 20th of May there were 31017 deaths in Italy, all age attributed. Germany has similarly recorded age for all deaths and is in fully updated at 23rd of March (Note: German data are reported in German from Robert Kock Institute; their age stratified data is reported in form of a plot. I sidelined retrieving data from the plot by accessing Wikipedia data which looks appropriate.) We can take the age pyramid of the two countries and rescale appropriately to check how Italian and German deaths would have appeared in Spain by accounting for demographic differences.

```r
buildDataset <- function( deaths.9, pyram.17, def.pyram.9)  {
    pyram.9 <- get.10y.from.5y(pyram.17)
    deaths.rescaled.9 <- (deaths.9/pyram.9)*def.pyram.9
    deaths.rescaled.9 <- deaths.rescaled.9*27650/sum(deaths.rescaled.9)
    return (deaths.rescaled.9)
}
# Rescaling for Spanish data (no rescale needed here!)
cases.deaths.rescaled.9 <- cases.deaths.9

# Loading and rescaling of Italian data
ITA <- c(4, 0, 14, 61, 268, 1101, 3219, 8447, 12691+5212)
ita.age.pyram.17 <- as.matrix( read.table("Data/Italy_5km_stats.dat",
                                          header=FALSE) )[,3]
ita.deaths.rescaled.9 <- buildDataset( ITA, ita.age.pyram.17, age.pyram.9 )

# Loading and rescaling of German data
GER <- c(1, 2, 8, 20, 62, 279, 761, 1844, 3689+1496+50)
ger.age.pyram.17 <- as.matrix( read.table("Data/Germany_5km_stats.dat",
                                          header=FALSE))[,3]
ger.deaths.rescaled.9 <- buildDataset( GER, ger.age.pyram.17, age.pyram.9 )

# Datasets and Plots
ver.projections.9 <-
    as.vector((age.pyram.9*IFR)*sum(cases.deaths.rescaled.9)/sum(age.pyram.9*IFR))
df1 <- data.frame( age=seq(0,8*10,10), ita=as.vector(ita.deaths.rescaled.9),
                   ger=as.vector(ger.deaths.rescaled.9),
                   esp=cases.deaths.rescaled.9,
                   ver=ver.projections.9)
tail(df1, n=10)
```

```
##   age          ita          ger          esp          ver
```

```
## 1   0     4.676917     3.815410     2.882310     3.478736
## 2  10     0.000000     8.544874     7.205775    16.717433
## 3  20    14.227736    25.248381    33.146565    71.174361
## 4  30    66.761145    68.737791    90.792766   239.274128
## 5  40   304.152275   304.233308   289.672157   610.868171
## 6  50  1084.241525   947.347881   880.545710  2049.188886
## 7  60  3119.473622  2536.680508  2442.757740  5097.205014
## 8  70  7556.523628  6376.405180  6675.430001  8444.863393
## 9  80 15499.943151 17378.986666 17227.566976 11117.229877
```

```r
print(colSums(df1))
```

```
##   age   ita   ger   esp   ver
##   360 27650 27650 27650 27650
```

```r
max.y = max(df1)
plot(c(0,80), c(0,max.y), type="n", xlab="Age", ylab="Number of deaths",
     main="Projected deaths according to Spanish age pyramid")
lines(df1$age, df1$esp, col="red", type='l')
lines(df1$age, df1$ita, col="blue")
lines(df1$age, df1$ger, col="orange")
lines(df1$age, df1$ver, col="darkgreen", lw=2, lt=2)
legend(0, 16000, c("Spain", "Italy", "Germany", "Verity pred."),
       col=c("red","blue","orange","darkgreen"), lty=c(1,1,1,2), lw=c(1,1,1,2))
```

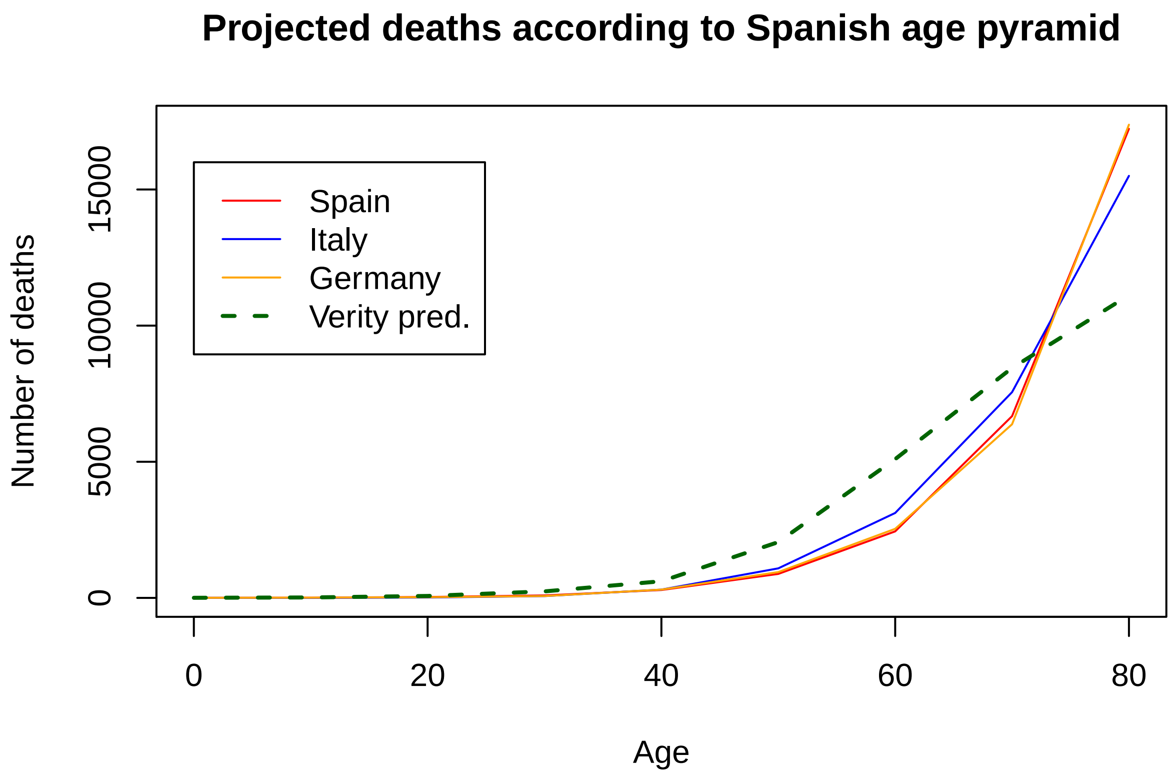


It is evident that after rescaling, the Spanish, German and Italian sets have more similarity than the predictions from the Verity IFR. It also appears interesting that German and Spanish data sets are almost superimposing, suggesting that the Spanish dataset is sufficiently robust. This also reassures on the fact that initially we proportionally redistributed, over all the age groups, the difference between the 27650 cases and the 19186 cases that had age information. However, it would be important to compare with additional countries. We add here a similar comparison with the additional sets from Portugal, Switzerland and Canada.

```
POR <- c(0, 0, 1, 0, 15, 41, 118, 258, 384+513)
por.age.pyram.17 <- as.matrix( read.table("Data/Portugal_5km_stats.dat",
                                          header=FALSE) )[,3]
por.deaths.rescaled.9 <- buildDataset( POR, por.age.pyram.17, age.pyram.9 )

CHZ <- c(0, 0, 0, 5, 4, 34, 114, 320, 1084)
chz.age.pyram.17 <- as.matrix( read.table("Data/Switzerland_5km_stats.dat",
                                          header=FALSE) )[,3]
chz.deaths.rescaled.9 <- buildDataset( CHZ, chz.age.pyram.17, age.pyram.9 )

# Loading and rescaling of Italian data
CAN <- c(0, 0, 3, 10, 36, 116, 382, 949, 3982+2153+1828)
can.age.pyram.17 <- c(1929522, 1993849, 1905977, 2067651, 2495075, 2575863,
                      2579123, 2501565, 2379067, 2363496, 2664738,
                      2725718, 2363696, 2019832, 1538354, 1056484, 817778)
can.deaths.rescaled.9 <- buildDataset( CAN, can.age.pyram.17, age.pyram.9 )

df2 <- data.frame( age=seq(0,8*10,10), por=as.vector(por.deaths.rescaled.9),
                   chz=as.vector(chz.deaths.rescaled.9),
                   can=as.vector(can.deaths.rescaled.9))

df0 <- merge(df1, df2, by.x='age', by.y='age')
```

Plots follow

```
max.y=max(df0)
plot(c(0,80), c(0, max.y), type='n', xlab="Age", ylab="Number of deaths",
     main = "Projected deaths according to Spanish age pyramid")
lines(df0$age, df0$esp, type='l', col="red")
lines(df0$age, df0$ita, col="blue")
lines(df0$age, df0$ger, col="orange")
lines(df0$age, df0$por, col="violet")
lines(df0$age, df0$chz, col="pink")
lines(df0$age, df0$can, col="black")
lines(df0$age, df0$ver, col="darkgreen", lw=2, lty=2)
legend(0,25000, c("Spain", "Italy", "Germany", "Portugal", "Switzerland",
                  "Canada", "Verity pred."), col=c("red", "blue",
                  "orange", "violet", "pink", "black", "darkgreen"),
       lty=c(1,1,1,1,1,1,2), lw=c(1,1,1,1,1,1,2))
```

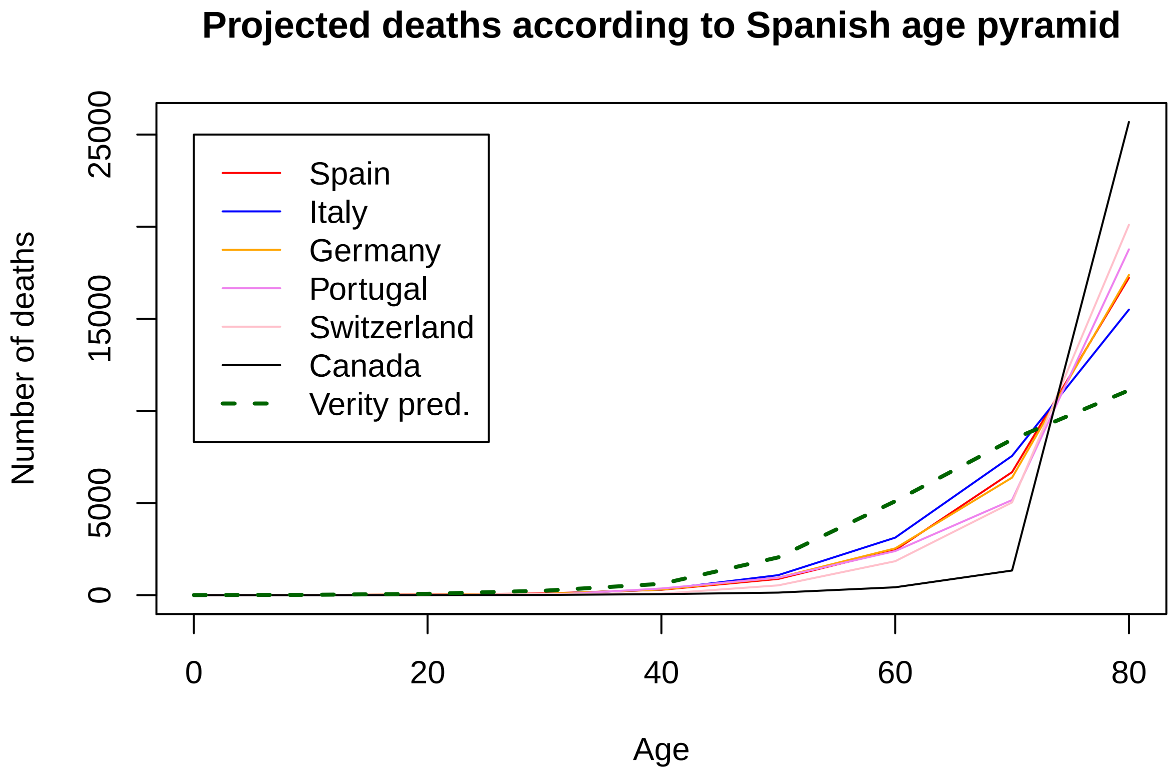


With respect to the other countries, Canada is remarkably different. However, cases are here renormalized so that we are taking age stratified cases in one country and compare with the Spanish case by accounting for demography and matching the total count number.

According to Wikipedia, the majority of deaths in earderly in Canada occurred in long-term care home with "factors such as outside visitors, communal living spaces, and staff being transferred among multiple facilities, as particular vulnerabilities.". Estimates suggest that "as many of 79 percent of Canada's COVID-19 fatalities occurred in long-term care homes". So it is a problem common to many countries that was particularly exacerbated in Canada. Hence, do the curves resamble each other if we exclude the 80+ category? We can normalize up to the 70s and observe how close the curves are:

```
makePlot <- function(nn)  {
    colours=c("red","blue", "orange","violet", "pink", "black", "darkgreen")
    max.y = 8000
    plot(c(0,(nn-1)*10), c(0,max.y), type="n", xlab="Age", ylab="Number of deaths",
         main = "Projected deaths renormalized to match the total of cases up to 70 y.o.")
    lines(df0$age[1:nn], df0$esp[1:nn]/sum(df0$esp[1:nn])*sum(df0$esp[1:nn]), type='l', col=colours[1])
    lines(df0$age[1:nn], df0$ita[1:nn]/sum(df0$ita[1:nn])*sum(df0$esp[1:nn]), type='l', col=colours[2])
    lines(df0$age[1:nn], df0$ger[1:nn]/sum(df0$ger[1:nn])*sum(df0$esp[1:nn]), type='l', col=colours[3])
    lines(df0$age[1:nn], df0$por[1:nn]/sum(df0$por[1:nn])*sum(df0$esp[1:nn]), type='l', col=colours[4])
    lines(df0$age[1:nn], df0$chz[1:nn]/sum(df0$chz[1:nn])*sum(df0$esp[1:nn]), type='l', col=colours[5])
    lines(df0$age[1:nn], df0$can[1:nn]/sum(df0$can[1:nn])*sum(df0$esp[1:nn]), type='l', col=colours[6])
    lines(df0$age[1:nn], df0$ver[1:nn]/sum(df0$ver[1:nn])*sum(df0$esp[1:nn]), type='l', col=colours[7],
          lw=2, lty=2)
    legend(0, 8000, c("Spain","Italy", "Germany", "Portugal", "Switzerland",
                      "Canada", "Verity pred."), col=colours,
           lty=c(1,1,1,1,1,1,2), lw=c(1,1,1,1,1,1,2))
```

```
}

makePlot(8)
```

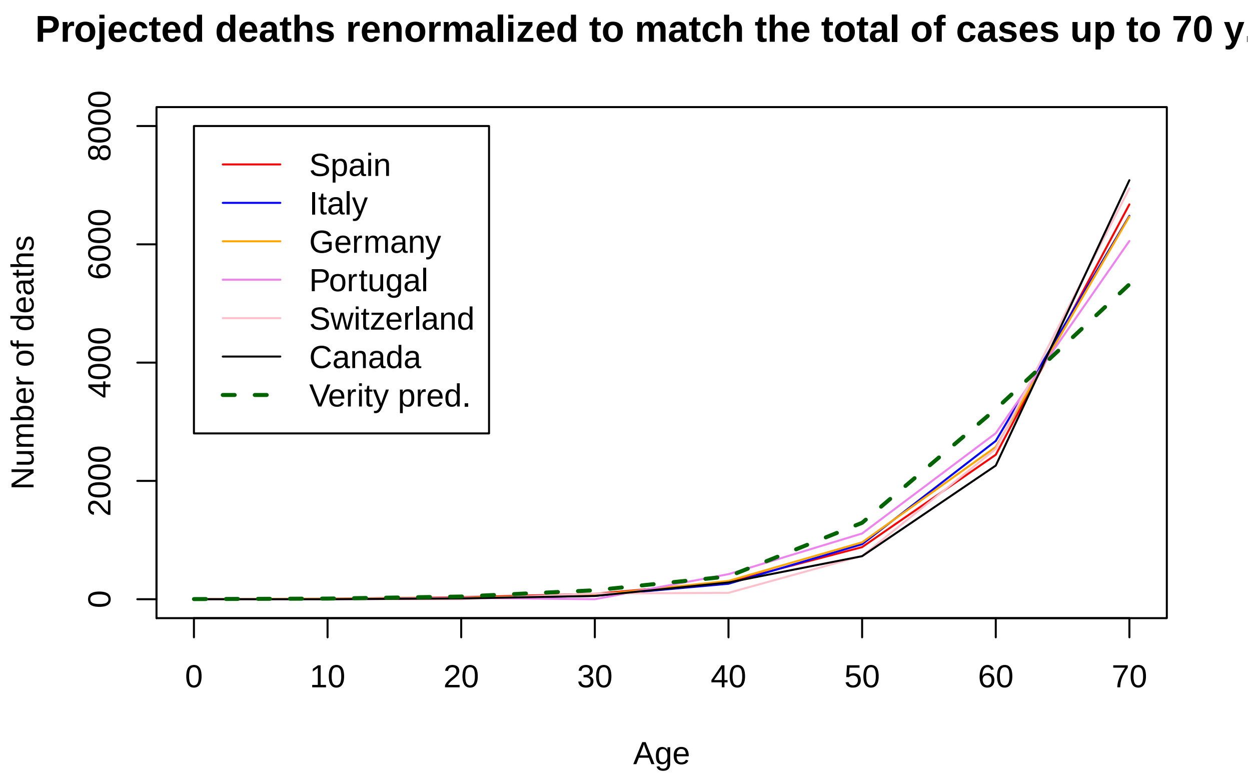


The plot still shows some variability in death incidence, but such variability is less dramatic and probably determined by stochastic effects and difference in age mixing. The Verity predictions are still somehow off. With this, we can thus assume that the the assumptions made about Spanish data are fine and we can save the rate to an external file.

```
#dat$s2h <- dat$s2h * 27650/19186.0 # <- C++ prgrammer idiosyncracy
#dat$s2c <- dat$s2c * 27650/19186.0
#dat$srate <- dat$srate * 27650.0/19186

print( paste('Check: 27650 =',sum(cases.total.9 * (dat$s2h*dat$h2d + dat$s2c*dat$c2d)) ) )
```

```
## [1] "Check: 27650 = 0"
```

```
write.csv(dat, file="Output/rates.csv", row.names=FALSE)
tail( dat, n=18 )
```

```
##   age       arate       a2s         a2c        s2d        c2d
## 1   0 0.006781274 0.2736602 0.044469783 0.01200963 0.00000000
## 2  10 0.007714451 0.1551619 0.014050092 0.02836919 0.00000000
## 3  20 0.054929514 0.1031127 0.006611693 0.02388081 0.00000000
## 4  30 0.081933888 0.1562348 0.012103008 0.01875757 0.08922351
## 5  40 0.073425664 0.2255870 0.021061619 0.02471234 0.12675967
## 6  50 0.091024028 0.2924008 0.036500444 0.05000437 0.16315049
## 7  60 0.093547221 0.4286962 0.073748545 0.11099852 0.31876460
```

```
## 8  70 0.107017749 0.5712480 0.069691459 0.30707026 0.43542543
## 9  80 0.289321537 0.4446118 0.006956023 0.68470426 0.63634337
```

```
sum(cases.deaths.9/(dat$arate*(dat$a2s*dat$s2d+dat$a2c*dat$c2d)))/sum(age.pyram.9)
```

```
## [1] 0.05081475
```

```
sum((spanish.estimate.count.9+cases.deaths.9)*dat$arate)
```

```
## [1] 239125
```

```
sum((spanish.estimate.count.9+cases.deaths.9)*dat$arate*(dat$a2s*dat$s2d+dat$a2c*dat$c2d))
```

```
## [1] 27650
```

```
sum(cases.deaths.9/(dat$arate*(dat$a2s*dat$s2d+dat$a2c*dat$c2d))-cases.deaths.9)
```

```
## [1] 2647143
```

```
sum(spanish.estimate.count.9)
```

```
## [1] 2647143
```

We can assume that all rates up to 70 y.o. are reasonably reliable, whilst death rates for 80+ is less reliable and strongly dependent on social factors.

### Estimating number of infected worldwide

We could use the number of deaths and the serological survey to estimate the fraction of the population infected by COVID-19. For instance Italy:

```
ita.age.pyram.9 <- get.10y.from.5y( ita.age.pyram.17 )
ita.est.9 <- ita.age.pyram.9*dat$srate*(dat$s2h*dat$h2d+dat$s2c*dat$c2d)*9
print(ita.est.9)
```

```
## numeric(0)
```

```
print(ITA)
```

```
## [1]     4     0    14    61   268  1101  3219  8447 17903
```

```
#sum(GER) / sum((get.10y.from.5y(ger.age.pyram.17)*sympt.rate.9*
#            (symp_to_hosp.9*hosp_to_death.9+symp_to_ICUs.9*critic_to_death.9)))*100.0
```

This of course works if deaths are registered similarly among nations. In addition, to apply this in general, we would need information on the distribution of cases by age for each country. This is not available for all countries, so we adopt a very rough approach by assuming that the cases do follow the expected distribution. This would not be correct, for instance, for Canada, and is likely to be a bit far-fetched. However, if we do insist in this approach, we can obtain an estimate of the fraction of the population who would result positive to a serological study in each country. We can use the JHSSE data set for the number of deaths (just to be sure I use data corresponding to 2 days earlier than the last day reported) and the UN estimates of population pyramids.

Read global datasets on deaths and population pyramids

```
library(dplyr)
```

```
##
## Attaching package: 'dplyr'
```

```
## The following objects are masked from 'package:stats':
##
##     filter, lag
## The following objects are masked from 'package:base':
##
##     intersect, setdiff, setequal, union
```

```
SELECTED.DATE <- 'X5.31.20'
deaths <- read.csv("Data/time_series_covid19_deaths_global.csv", header=TRUE,
                   stringsAsFactors = FALSE)
pops <- read.csv("Data/WPP2019_POP_F07_1_POPULATION_BY_AGE_BOTH_SEXES.csv",
                 header=TRUE, skip=16, stringsAsFactors = FALSE)
```

Perform some modification to match country names and exclude non-matching/missing countries.

```
d.countries <- unique(deaths$Country.Region)

names <- colnames(pops)
names[3] <- "Country"
colnames(pops) <- names
# Reset some country names (incomplete)
pops[pops$Country == "Iran (Islamic Republic of)",]$Country <- "Iran"
pops[pops$Country == "Russian Federation",]$Country <- "Russia"
pops[pops$Country == "Republic of Korea",]$Country <- "South Korea"
pops[pops$Country == "United States of America",]$Country <- "US"
pops[pops$Country == "Syrian Arab Republic",]$Country <- "Syria"

p.countries <- unique(pops$Country)
commonCountries <- d.countries[(d.countries %in% p.countries)]
deaths <- select( deaths[which(deaths$Country.Region %in% commonCountries),],
                  Country.Region, Province.State, SELECTED.DATE )
```

Group countries that are detailed in terms of provinces/territories (for some countries this is incorrect, i.e. the UK, but the difference is quite small) and merge the two data sets

```
deaths <- summarise_at( group_by(deaths, Country.Region), vars(SELECTED.DATE), sum)
pops <- pops[which(pops$Country %in% commonCountries),]
full <- merge(pops, deaths, by.x='Country', by.y='Country.Region', all=TRUE)
full <- full[full$Reference.date..as.of.1.July. == 2020,]
```

Build estimates by distributing deaths to age groups and reverting transformation to positives using Spanish survey data. Also add projected deaths for 90% infected population

```
format.rate <- function(x)  {return(floor(10000*x+0.5)/100.0)}
transRate.9 <- dat$arate*(dat$a2s*dat$s2d+dat$a2c*dat$c2d)
globalEstimates <- data.frame( Country=as.character(),
                     positive=as.numeric(), stringsAsFactors = FALSE )
distributeCases <- function( total, distr )  {
    normdistr <- distr / sum(distr)
    return(total*normdistr)
}

getEstimates <- function( row ) {
    pyramid.21 <- as.numeric(gsub(" ", "", row[9:29]))
    pyramid.17 <- pyramid.21[1:17]*1000 # in thousands
    pyramid.17[17] <- pyramid.17[17] + sum(pyramid.21[18:21])*1000 # idem
```

```r
    pyramid.9  <- get.10y.from.5y( pyramid.17 )
    t.deaths <- as.numeric(gsub(" ","", row[SELECTED.DATE]))
    d.deaths <- distributeCases(t.deaths, transRate.9*pyramid.9)
    globalEstimates <<- rbind(globalEstimates, data.frame( Country=row$Country,
               Positives=format.rate(sum(d.deaths / transRate.9-d.deaths) /
                                   sum(pyramid.9)),
               Deaths=t.deaths,
               Projection=sum(pyramid.9*transRate.9*0.9) ))
}
dummy <- by(full, 1:nrow(full), getEstimates) # This avoids output
globalEstimates <- globalEstimates[order(-globalEstimates$Positives),]
print(globalEstimates)
```

```
##                              Country Positives Deaths   Projection
## 16                           Belgium     10.06   9467   84028.2131
## 157                   United Kingdom      7.47  38571  461109.7097
## 140                            Spain      6.62  27127  365345.3562
## 44                           Ecuador      6.60   3358   45684.9011
## 74                           Ireland      6.19   1652   23883.6817
## 144                           Sweden      5.41   4395   72509.2250
## 76                             Italy      5.39  33415  552646.2487
## 54                            France      5.07  28805  506604.5915
## 159                               US      4.97 104381 1878849.5660
## 128            Sao Tome and Principe      4.74     12     227.3916
## 106                      Netherlands      4.52   5975  117990.2638
## 118                             Peru      4.19   4506   96526.5831
## 72                              Iran      3.92   7797  178389.5068
## 82                            Kuwait      3.90    212    4886.5718
## 22                            Brazil      3.85  29314  683130.4687
## 156             United Arab Emirates      3.39    264    6995.8362
## 90                        Luxembourg      2.98    110    3302.2287
## 145                      Switzerland      2.81   1920   61063.9057
## 29                            Canada      2.80   7374  235780.8320
## 99                            Mexico      2.65   9930  336260.3427
## 115                           Panama      2.29    336   13157.6589
## 153                           Turkey      1.63   4540  249372.7088
## 43                Dominican Republic      1.58    502   28593.7015
## 122                            Qatar      1.57     38    2180.8110
## 42                          Djibouti      1.48     24    1458.2015
## 121                         Portugal      1.45   1410   86660.3178
## 111                  North Macedonia      1.32    133    9006.6181
## 41                           Denmark      1.30    574   39502.1021
## 32                             Chile      1.18   1054   80317.6990
## 67                          Honduras      1.09    212   17447.9216
## 11                           Bahamas      1.07     11     921.3806
## 58                           Germany      1.06   8540  717344.1130
## 129                     Saudi Arabia      1.04    503   43504.0890
## 47                 Equatorial Guinea      0.96     12    1120.1735
## 12                           Bahrain      0.95     19    1807.0085
## 7                            Armenia      0.92    131   12799.6204
## 9                            Austria      0.92    668   64566.6010
## 123                          Romania      0.89   1266  126910.1909
## 113                             Oman      0.85     49    5172.3079
## 5                Antigua and Barbuda      0.78      3     344.0338
```

```
## 28                  Cameroon      0.77    191   22262.2749
## 68                   Hungary      0.75    526   62795.2154
## 1                Afghanistan      0.73    257   31806.2767
## 20    Bosnia and Herzegovina      0.73    153   18753.0394
## 75                    Israel      0.69    285   36783.4472
## 53                   Finland      0.67    320   42647.3042
## 112                   Norway      0.64    236   32716.8851
## 136                 Slovenia      0.64    108   15113.8949
## 138             South Africa      0.62    683   98956.6706
## 49                   Estonia      0.61     68    9952.5330
## 94                  Maldives      0.60      5     749.4892
## 3                    Algeria      0.59    653   98727.1587
## 55                     Gabon      0.59     17    2574.2535
## 65                    Guyana      0.57     12    1882.1575
## 133             Sierra Leone      0.57     46    7278.3752
## 34                  Colombia      0.53    916  154533.5836
## 124                   Russia      0.53   4693  787887.5900
## 45                     Egypt      0.51    959  169017.0994
## 69                   Iceland      0.51     10    1758.5165
## 137                  Somalia      0.51     78   13872.7515
## 142                    Sudan      0.51    286   50525.7420
## 31                      Chad      0.46     65   12791.2291
## 87                   Liberia      0.46     27    5274.2592
## 95                      Mali      0.46     77   15156.0193
## 119              Philippines      0.45    957  191634.4717
## 64             Guinea-Bissau      0.44      8    1639.7769
## 73                      Iraq      0.43    205   43354.8325
## 97                Mauritania      0.43     23    4795.9751
## 114                 Pakistan      0.43   1483  311657.8213
## 40                   Czechia      0.42    320   67953.8181
## 131                   Serbia      0.42    243   51754.6764
## 15                   Belarus      0.41    235   51064.0250
## 147               Tajikistan      0.40     47   10499.9247
## 120                   Poland      0.39   1064  241849.1309
## 14                  Barbados      0.36      7    1751.3288
## 26                Cabo Verde      0.34      4    1059.6345
## 109                    Niger      0.32     64   18224.4343
## 62                 Guatemala      0.31    108   31543.6743
## 24              Burkina Faso      0.30     53   15667.2667
## 37                   Croatia      0.30    103   30894.8862
## 89                 Lithuania      0.29     70   21420.2315
## 6                  Argentina      0.27    539  176271.0929
## 71                 Indonesia      0.27   1613  539213.9839
## 17                    Belize      0.26      2     689.0354
## 23                  Bulgaria      0.26    140   48399.5303
## 39                    Cyprus      0.26     17    5846.1141
## 162                    Yemen      0.26     80   27265.8683
## 96                     Malta      0.25      9    3238.1931
## 101               Montenegro      0.25      9    3183.3153
## 155                  Ukraine      0.25    708  254724.7653
## 10                Azerbaijan      0.24     63   23980.2255
## 108                Nicaragua      0.24     35   12956.3072
## 130                  Senegal      0.24     42   15991.7712
## 2                    Albania      0.22     33   13693.6297
```

```
## 35                    Comoros      0.21     2      850.4859
## 46                El Salvador      0.21    46    19479.3128
## 66                      Haiti      0.21    44    19067.9902
## 102                   Morocco      0.21   205    87071.7666
## 13                 Bangladesh      0.19   650   312377.3310
## 63                     Guinea      0.18    23    11313.0115
## 98                  Mauritius      0.18    10     4862.5796
## 60                     Greece      0.17   175    93656.4257
## 70                      India      0.17  5408 2780151.0603
## 110                   Nigeria      0.16   287   164385.2872
## 150                      Togo      0.16    13     7194.5072
## 84                     Latvia      0.15    24    14106.4251
## 161           Western Sahara      0.15     1      595.7081
## 83                 Kyrgyzstan      0.14    16    10590.5640
## 85                    Lebanon      0.14    27    17491.6671
## 93                   Malaysia      0.14   115    73222.0499
## 151       Trinidad and Tobago      0.14     8     4954.4870
## 81                      Kenya      0.13    64    43497.2782
## 152                   Tunisia      0.13    48    34349.4938
## 38                       Cuba      0.12    83    62313.9371
## 50                   Eswatini      0.12     2     1472.6063
## 59                      Ghana      0.11    36    30717.5481
## 158                   Uruguay      0.10    22    19838.1527
## 77                    Jamaica      0.09     9     9275.2909
## 134                 Singapore      0.09    23    23885.0243
## 135                  Slovakia      0.09    28    28486.0076
## 33                      China      0.08  4638 5056339.1960
## 139               South Sudan      0.08    10    11881.6506
## 80                 Kazakhstan      0.07    40    50064.9220
## 107               New Zealand      0.07    22    26935.4876
## 8                   Australia      0.06   103   146013.9134
## 78                      Japan      0.06   898 1372976.6915
## 79                     Jordan      0.06     9    13708.3650
## 117                  Paraguay      0.06    11    15764.6236
## 143                  Suriname      0.06     1     1412.3891
## 36                 Costa Rica      0.05    10    17947.0979
## 56                     Gambia      0.05     1     1888.7926
## 57                    Georgia      0.05    12    21070.0245
## 163                    Zambia      0.05     7    13025.5972
## 30   Central African Republic      0.04     2     4162.8556
## 88                      Libya      0.04     5    10787.8937
## 21                   Botswana      0.03     1     3129.2073
## 160                Uzbekistan      0.03    15    53840.7158
## 18                      Benin      0.02     3    12586.2127
## 91                 Madagascar      0.02     6    27456.5731
## 92                     Malawi      0.02     4    15933.7040
## 146                     Syria      0.02     5    27923.6139
## 148                  Thailand      0.02    57   305088.2975
## 164                  Zimbabwe      0.02     4    14425.2887
## 4                      Angola      0.01     4    24021.6951
## 25                    Burundi      0.01     1     8922.2868
## 51                   Ethiopia      0.01    11   128291.0324
## 103                Mozambique      0.01     2    27581.6763
## 105                     Nepal      0.01     8    50535.2896
```

```
## 125                          Rwanda      0.01       1    12426.1099
## 141                       Sri Lanka      0.01      10    70946.5282
## 19                           Bhutan      0.00       0     1676.0381
## 27                         Cambodia      0.00       0    24104.2024
## 48                          Eritrea      0.00       0     4836.3388
## 52                             Fiji      0.00       0     1475.8306
## 61                          Grenada      0.00       0      355.6162
## 86                          Lesotho      0.00       0     3358.3195
## 100                        Mongolia      0.00       0     4847.1444
## 104                         Namibia      0.00       0     3033.6623
## 116                Papua New Guinea      0.00       0     9336.9268
## 126                     Saint Lucia      0.00       0      630.4370
## 127 Saint Vincent and the Grenadines      0.00       0      353.2166
## 132                      Seychelles      0.00       0      243.5598
## 149                     Timor-Leste      0.00       0     1733.2016
## 154                          Uganda      0.00       0    29103.8576
```

Note that we are using the same approach for Spain, and the resulting percentage is higher than the one measured by the survey. This is a direct consequence of the fact that the implied distribution of cases is not the true one (checked, not shown). Not sure how to associate an error (bootstrapping?).

Summing up all projections we have the expected number of worldwide deaths.

```
print(paste('Expected number of worldwide deaths is:',
            floor(sum(globalEstimates$Projection))/1000000,'millions'))
```

```
## [1] "Expected number of worldwide deaths is: 22.64082 millions"
```

### References


Spanish survey [ https://www.mscbs.gob.es/gabinetePrensa/notaPrensa/pdf/13.05130520204528614.pdf ]

Spanish cases at 17.05.2020 [ https://www.mscbs.gob.es/profesionales/saludPublica/ccayes/alertasActual/nCov-China/documentos/Actualizacion_108_COVID-19.pdf ]

Italian ICU study [ https://jamanetwork.com/journals/jama/fullarticle/2764365 ]

Estimate of Rt in Lombardy [ https://www.medrxiv.org/content/10.1101/2020.04.08.20056861v1 ]

JHSSE data set data

Population estimates United Nations, Department of Economic and Social Affairs, Population Division (2019). World Population Prospects 2019, Online Edition. Rev. 1.
